# Single-cell RNA-sequencing of PBMCs from SAVI patients reveals disease-associated monocytes with elevated integrated stress response

**DOI:** 10.1101/2023.04.25.23288913

**Authors:** Camille de Cevins, Laure Delage, Maxime Batignes, Quentin Riller, Marine Luka, Anne Remaury, Boris Sorin, Tinhinane Fali, Cécile Masson, Bénédicte Hoareau, Catherine Meunier, Mélanie Parisot, Mohammed Zarhrate, Brieuc P. Pérot, Víctor García-Paredes, Francesco Carbone, Luc Canard, Charlotte Boussard, Etienne Crickx, Jean-Claude Guillemot, Marie-Louise Frémond, Bénédicte Neven, Galina Boldina, Franck Augé, Fischer Alain, Michel Didier, Frédéric Rieux-Laucat, Mickael M. Ménager

## Abstract

Gain-of-function mutations in *STING1*, which encodes the Stimulator of Interferon Gene (STING), result in a severe autoinflammatory disease termed STING-associated vasculopathy with onset in infancy (SAVI). Although elevated type I interferon (IFN) production is thought to be the leading cause of the symptoms observed in patients, STING can induce a set of pathways, which roles in the onset and severity of SAVI, remain to be elucidated. To address this point, we compared a single-cell RNA sequencing (scRNA-seq) dataset of peripheral blood mononuclear cells (PBMCs) from SAVI patients to a dataset of healthy PBMCs treated with recombinant IFN-β. We revealed a loss of mucosal associated invariant T cells and CD56^bright^ natural killer cells in SAVI patients, not observed in IFN-β-treated PBMC. Patients’ T cells present markers of early activation, associated with markers of senescence and apoptosis. Inferring cell-to-cell communication from scRNA-seq predicted monocytes as potential drivers of this T cell phenotype. Furthermore, scRNA-seq clustering identified a patient-specific subset of monocytes, expressing a strong integrated stress response (ISR), and high *CCL3*, *CCL4* and *IL-6*. It also pinpointed to a patient with lower ISR, allowing us to identify a secondary mutation in PERK, recently shown to be activated by STING to trigger the ISR. Finally, based on the identification of this patient-specific subset of monocytes and the exploration of IFN-β stimulated PBMCs from healthy donors, we developed a strategy to propose a transcriptomic signature specific of STING activation and independent of type I IFN response. Altogether, these results provide a deeper understanding of SAVI at the cellular and molecular levels.

## Introduction

Stimulator of Interferon genes (STING)-associated vasculopathy with onset in infancy (SAVI) is a rare monogenic disease characterized by uncontrolled production of type I interferons (IFNs)^1, 2^. Type I IFNs are the first line of defense against pathogenic infection. They lead to a robust activation of the immune system and increase the effector functions of immune cells while triggering apoptosis of infected cells. However, type I IFNs are highly potent cytokines that can become detrimental when uncontrolled. SAVI is a type I interferonopathy that usually appears during the first few years of life and is characterized by severe pulmonary and cutaneous manifestations^3^. SAVI is caused by gain-of-function mutations in *STING1,* which codes for STING, a key component of innate immunity, capable of eliciting a potent innate immune response. To date, about 100 cases of SAVI have been reported^4^.

The STING pathway acts as a sensor of cytoplasmic nucleic acids, which can represent a potential danger signal. Upon sensing of cytoplasmic nucleic acids, STING is activated and changes conformation to traffic from the endoplasmic reticulum (ER) to the Golgi apparatus^5, 6^. There, STING triggers the phosphorylation of IRF3 and NF-κB, leading to the expression of type I IFNs and other proinflammatory cytokines^7–9^. Other molecular pathways can also be triggered by STING, such as ER stress and senescence^10, 11^. Moreover, STING can induce cell type-specific programs, such as apoptosis, which is induced in T cells but not in macrophages or dendritic cells^12^.

In SAVI, STING is constitutively activated and present at the Golgi apparatus^1, 13^. SAVI is associated with T cell lymphopenia, and an imbalance between memory and naïve CD8^+^ T cells, with decreased percentages of effector and memory T cells^3, 14^. A low number of circulating NK cells has also been reported in some patients^15^. At the molecular levels, IFN- α/β and IFN-stimulated genes (ISGs) are consistently upregulated in the plasma of all patients^1–3^. Patients have elevated inflammatory cytokines plasma concentrations, including Tumor Necrosis Factor-α (TNF-α) and Interleukin-6 (IL-6)^16^, suggesting the involvement of the NF- κB pathway. Patients’ vascular endothelial cells also have increased expression of genes involved in apoptosis, cell adhesion, and coagulation^2^. Many pathways are thought to be triggered by STING activation. Some of them, such as type I IFNs production and NF-κB activation, are upregulated in SAVI. Others, such as T cell activation or increased ER stress, have been demonstrated in murine models but have yet to be confirmed in SAVI patients^11^.

As uncontrolled production of type I IFNs is a hallmark of SAVI, treatment with Janus kinase (JAK)-inhibitors has been proposed to patients. These drugs block the signal transduction upon IFN-α/β binding to the IFN-α/β receptor (IFNAR). The JAK1/JAK2 inhibitors, Ruxolitinib and Baricitinib, are among the most administered in SAVI, with JAK1 acting downstream of IFNAR^3^. Tofacitinib, another JAK-inhibitor targeting JAK1 and JAK3 has also been used. These treatments lead to partial improvements of the symptoms^3, 17, 18^. However, in cases where the interstitial lung disease is too advanced, JAK-inhibitor treatment has proved inefficient^3, 17^ suggesting in SAVI, an alternative or combined role of STING activation, independent of type I IFN signaling. A deeper understanding of the pathways modulated in patients therefore appears crucial to propose alternative therapeutic targets/strategies. Here, we aim to identify pathways modulated in a cell-type-specific manner and address their dependency to primary type I IFN production.

The functional impairment of STING gain-of-function has mostly been assessed using *in vitro* cell lines or mouse models^11, 19, 20^. Others have used patients’ cells, but evaluated individual cell types^14^. Here, we propose to evaluate peripheral blood mononuclear cells (PBMCs) at the single cell level to gain a systemic overview of the transcriptomic dysregulations taking place in SAVI. This approach also aims to identify disease-associated cells, infer cell-cell communication, and compare the pathways specifically modulated in different immune cell types. Additionally, to decipher type I IFN-mediated signaling from other dysregulated pathways driven by STING constitutive activation, we propose to compare the primary response to IFN-β in PBMCs coming from healthy donors, to the pathways modulated in SAVI patients. We have therefore built two datasets, one with five SAVI patients before and after JAK-inhibitor treatment and seven healthy donors (SAVI dataset), and another with healthy PBMCs challenged with IFN-β at different timepoints (IFN-β dataset). Overall, we analyzed more than 220,000 cells at the single-cell transcriptomic level.

## Results

### Circulating effector lymphocytes are decreased in SAVI, including MAIT, γδ T cells, NK cells, effector CD4^+^ and CD8^+^ T cells

To better understand the molecular physiopathology of SAVI, we profiled at the single-cell transcriptomic level a dataset composed of PBMCs from five SAVI patients carrying different gain-of-function mutations in *STING1*, as well as seven healthy donors (CTRLs) (SAVI dataset) (Figures 1A and S1A). All patients were previously clinically described, as indicated in Figure S1A^1, 3, 15, 18, 21^. We collected blood samples before JAK-inhibitor initiation for all patients (SAVI group), and samples upon-JAK-inhibition for four of them (SAVI_treated group). These four patients were treated with the JAK-1/2 inhibitor Ruxolitinib for a duration of 1 to 52 months (4 years and 4 months), and one patient (P1) was later switched to the JAK- 1/3 inhibitor, Tofacitinib, for 2 months. For this patient (P1), a blood sample was collected upon each treatment.

**Figure 1:**
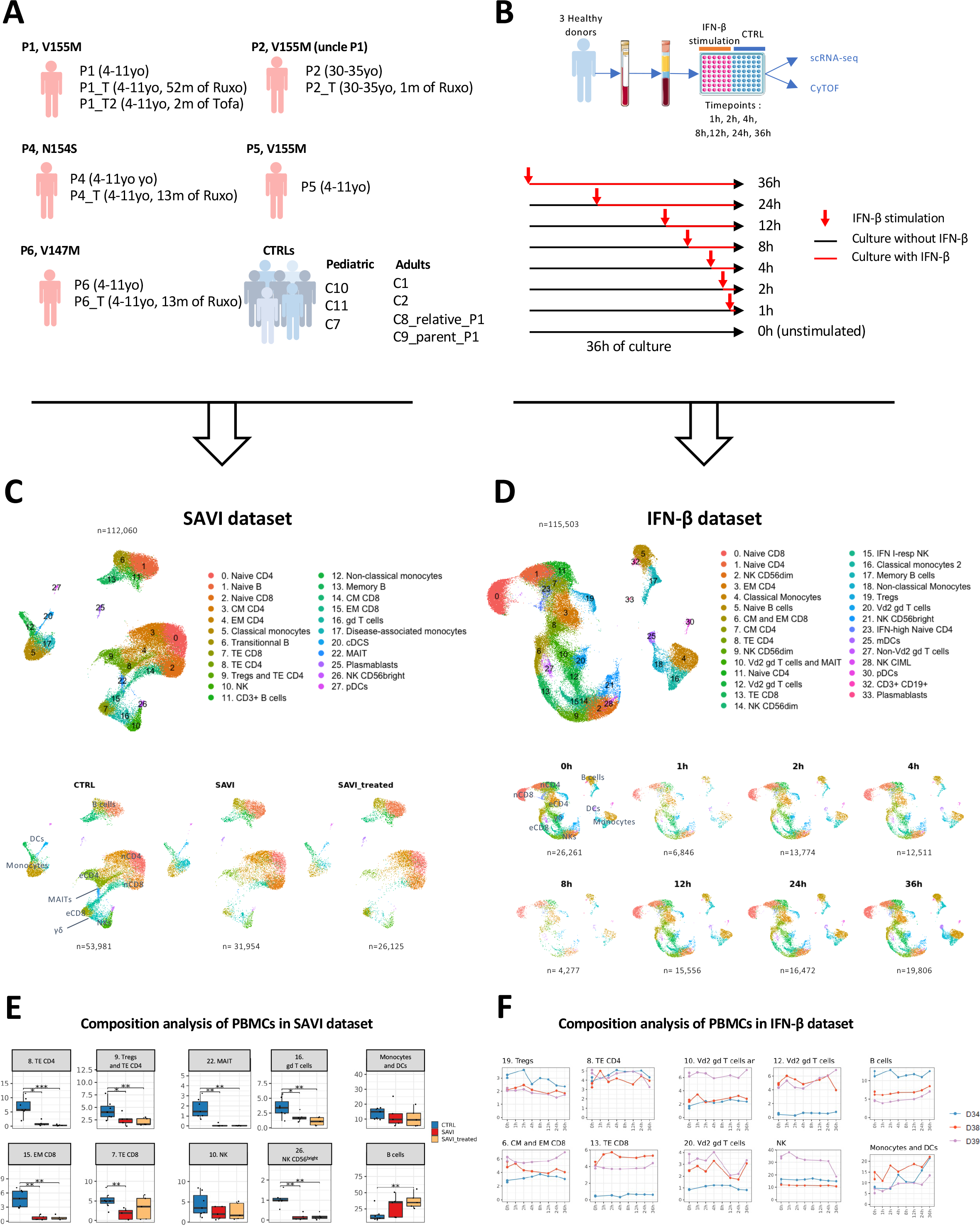
A scRNA-seq cohort of PBMCs from 5 SAVI patients shows loss of effector cells not replicated by challenging healthy PBMCs with IFN-β A. SAVI dataset: Cohort of 5 SAVI patients, with 3 different STING mutations (V155M, N154S, V147M), sampled before (SAVI) and under JAK-inhibitor treatment (SAVI_treated), and 7 healthy donors (CTRL). The age of each sample and the time of treatment is indicated (yo: years old; m: months). Ruxo: Ruxolitinib; Tofa: Tofacitinib B. IFN-β dataset: PBMCs from 3 healthy donors were cultured for 36h with addition of 1000 IU/mL of IFN-β at one of several timepoints C. UMAP and cell type assignment of 112,060 cells from the SAVI dataset (top) and UMAP of the SAVI dataset separated by group (CTRL, SAVI, SAVI_treated). The number of cells in each group is indicated below the UMAP (bottom). nCD4: naïve CD4; eCD4: effector CD4; nCD8: naïve CD8; eCD8: effector CD8. D. UMAP and cell type assignment of 115,503 cells from the IFN-β dataset (top) and UMAP of the IFN-β dataset separated by time of IFN-β stimulation (0h, 1h, 2h, 4h, 8h, 12h, 24h and 36h). The number of cells in each group is indicated below the UMAP (bottom). nCD4: naïve CD4; eCD4: effector CD4; nCD8: naïve CD8; eCD8: effector CD8. E. Boxplot of the proportion of PBMCs found in several clusters of the SAVI dataset. *p*- values are calculated by Kruskal-Wallis test for multiple comparisons, followed by a post hoc Dunn’s test. *(*p* < 0.05), **(*p* < 0.01), ***(*p* < 0.001). F. Evolution of the proportion of PBMCs found in several clusters in the IFN-β dataset over the time course of IFN-β stimulation.

In parallel, we aimed to better determine the contribution of type I IFN secretion to the transcriptomic profile of SAVI patients. To do so, we analyzed PBMCs from three healthy donors (donors D34, D38 and D39), following *in vitro* stimulation with human recombinant IFN-β at different timepoints, using the same single-cell approach (IFN-β dataset) (Figure 1B). The response to IFN-β stimulation was assessed in the scRNA-seq by following the kinetics of expression at the mRNA level of 6 ISGs commonly used to evaluate type I IFN response in patients^1^ (Figure S1B).

After quality control, integration, and unsupervised clustering of each single-cell transcriptomic dataset, we obtained 23 cell populations in the SAVI dataset and 29 in the IFN- β dataset (Figures 1C-D, S1C-F). We compared cell population proportions in SAVI patients to CTRLs (Figures 1C, 1E, S1G), and confirmed the previously described increase of naïve CD4^+^ and CD8^+^ T cells and decrease of their effector/memory counterpart^14^, as well as NK cells^22^ (particularly NK CD56^bright^ cells). In addition, we observed a drastic reduction in the proportion of mucosal associated invariant T cells (MAIT) and γδ T cells in SAVI patients (Figures 1C, 1E). We did not observe significant differences in cell type proportions before and under JAK-inhibitor treatment. These observations were subsequently validated by flow cytometry for the SAVI dataset and by Cytometry by time-of-flight (CyTOF) for the IFN-β dataset (Figures S1I-J). Of note, no overt decrease of effector T cells was observed in healthy PBMCs challenged *in vitro* with type I IFN for 36 hours *in vitro* (Figures 1D, 1F, S1H, S1J).

### Cells of SAVI patients display strong type I IFN response and NF-κB activation, alongside cell stress, death, and modulation of actin and integrin-related pathways

STING activation triggers NF-κB signaling and type I IFN production^23^. Therefore, we sought to evaluate these pathways in the immune cells of SAVI patients at the single-cell level. Response to type I IFNs was increased in all PBMC cell types in SAVI patients, with the strongest induction observed in monocytes and DCs (Figures 2A, S2A). This type I IFN response was partially corrected following JAK-inhibitor treatment, with the mean signature score being lower in treated patient compared to untreated, but remained higher than in CTRLs (Figure S2A). After *in vitro* stimulation with human recombinant IFN-β, a robust type I IFN response is measured, as soon as one hour of stimulation, peaking after 2h (Figures 2B, S2B). The strongest type I IFN response is also observed in monocytes and DCs of the IFN-β dataset, specially at later time points. In the SAVI dataset, the transcriptomic signature of NF-κB activation follows the pattern of type I IFN response (Figure 2C), with the strongest induction in monocytes and DCs of SAVI patients. Under JAK-inhibitor treatment, NF-κB activation was back to CTRL levels. In the IFN-β dataset, NF-κB activation was induced as soon as one hour post by IFN-β stimulation and continued to increase until 36h particularly in monocytes and DCs. (Figure 2D).

**Figure 2:**
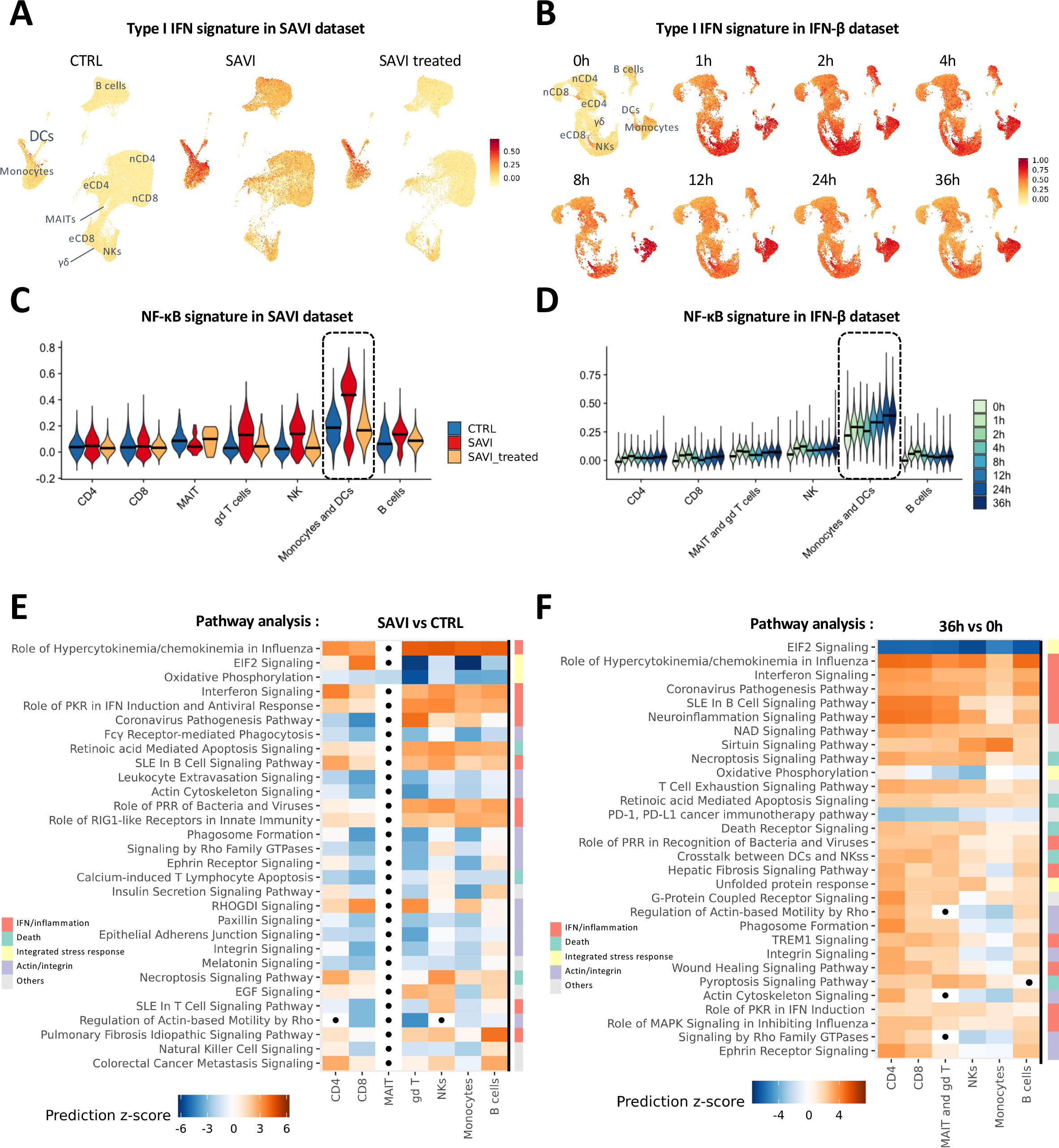
Type I IFN and NF-κB signature scores are elevated in SAVI patients A. Score in each cell of a type I IFN response signature of 272 genes, in the SAVI dataset. nCD4: naïve CD4; eCD4: effector CD4; nCD8: naïve CD8; eCD8: effector CD8; γδ: γδ T cells. B. Score in each cell of a type I IFN response signature of 272 genes, in the IFN-β dataset split by timepoints. nCD4: naïve CD4; eCD4: effector CD4; nCD8: naïve CD8; eCD8: effector CD8; γδ: γδ T cells. C. Violin plot of the score of an NF-κB activation signature of 200 genes, in the SAVI dataset. Dark lines indicate medians. In blue are the cells from the 7 CTRLs, in red the 5 SAVI patients, and in yellow the 5 SAVI patients treated with JAK-inhibitors. Dark lines indicate medians. D. Violin plot of the score of an NF-κB activation signature of 200 genes, in the IFN-β dataset. Dark lines indicate medians. Gradient of blues indicate time of IFN-β stimulation. E. Heatmap of the pathway enrichment analysis, performed in IPA, between SAVI and CTRL in each cell compartment. Dots indicate non-significant pathways (Bonferroni- Hochberg corrected p-values > 0.05). Side color bar indicate groups of pathways based on broader functions. Color scales indicate direction of prediction with orange for predicted activation and blue for predicted inhibition. F. Heatmap of the pathway enrichment analysis, performed in IPA, between healthy PBMCs from 3 different donors stimulated with IFN-β for 36h and unstimulated PBMCs in each cell compartment. Dots indicate non-significant pathways (Bonferroni-Hochberg corrected p-values > 0.05). Side color bar indicate groups of pathways based on broader functions. Color scales indicate direction of prediction with orange for predicted activation and blue for predicted inhibition.

To explore the dysregulated pathways in immune cells of SAVI patients, we used an approach based on differentially expressed genes (DEGs) between untreated SAVI and CTRLs in each major cell population, including CD4^+^ T cells, CD8^+^ T cells, γδ T cells, MAITs, NKs, B cells and monocytes and DCs. Pathway analysis was performed in QIAGEN Ingenuity Pathway Analysis (IPA) which predicts whether pathways are activated or inhibited based on the fold change of the members of the pathway, taking into account their activating or inhibiting role. We observed, in most cell types, an enrichment in pathways involved in response to IFNs and/or linked to inflammation (Figure 2E, supplementary file 1). In the SAVI dataset, the pathways related to IFN and inflammation were downregulated by JAK-inhibitor treatment but remained higher than in CTRLs (Figures S2A, S2C, supplementary file 1). IFN-related and inflammation-related pathways were also found to be activated in PBMCs treated with IFN-β for 36h (Figure 2F, supplementary file 2). Cell death pathways were enriched in both SAVI patients compared to CTRLs, and in PBMCs stimulated with IFN-β for 36h compared to unstimulated cells. JAK-inhibition also seemed to partially correct the induction of these cell death-related pathways (Figure S2C). In addition, we observed in SAVI an enrichment in pathways related to leukocyte trafficking and actin signaling. Interestingly, whereas actin/integrin-related pathways are predicted to be downregulated in all PBMCs in SAVI patients, these pathways are predicted to be upregulated in T cells and B cells, while inhibited in monocytes and NK cells in healthy PBMCs, 36h post‒IFN-β‒stimulation. (Figures 2E-F). Additionally, when looking both in PBMCs from SAVI patients and *in vitro* stimulated PBMCs, we observed a robust modulation of pathways that could be linked to the integrated stress response (ISR), with downregulation of eIF2 signaling and oxidative phosphorylation and an upregulation of the unfolded protein response (UPR). In the IFN-β dataset, this response was inhibited in a time-dependent manner, with the EIF2 signaling pathway being inhibited as soon as 2h, with the lowest prediction score at 36h. Meanwhile, the UPR was induced as soon as 4h with increasing levels of induction throughout the time course (Figure S2D, supplementary file 3).

### T lymphocytes of SAVI patients present a hyperactivated, senescent and apoptotic phenotype, associated with IFN-β signaling

Imbalance between naïve and effector T cells, along with proliferation defects, have previously been described in SAVI^14^. To improve our knowledge of transcriptomic dysregulations in circulating T cells from SAVI patients, we performed differential analysis between untreated SAVI patients and CTRLs in each cluster of the CD4^+^ and CD8^+^ T cell compartments (supplementary file 4). As expected, IPA revealed a strong enrichment of inflammatory response genes (Figure 3A). It also pointed to the enrichment of genes involved in actin and integrin-related pathways, and particularly the leukocyte extravasation signaling pathway (Figure 3A). Several other associated pathways, including RAC signaling and integrin signaling, were particularly decreased in effector cells. We hypothesized that this may reflect an impaired lymphocyte trafficking machinery, which was supported by the decrease of a lymphocyte trafficking signature score in T cells of SAVI patients, and particularly in effector T cells (Figure 3B). This signature score was also decreased after IFN-β challenge of PBMCs from healthy donors (Figure 3C), suggesting that type I IFNs may cause impaired T cell trafficking in SAVI patients.

**Figure 3:**
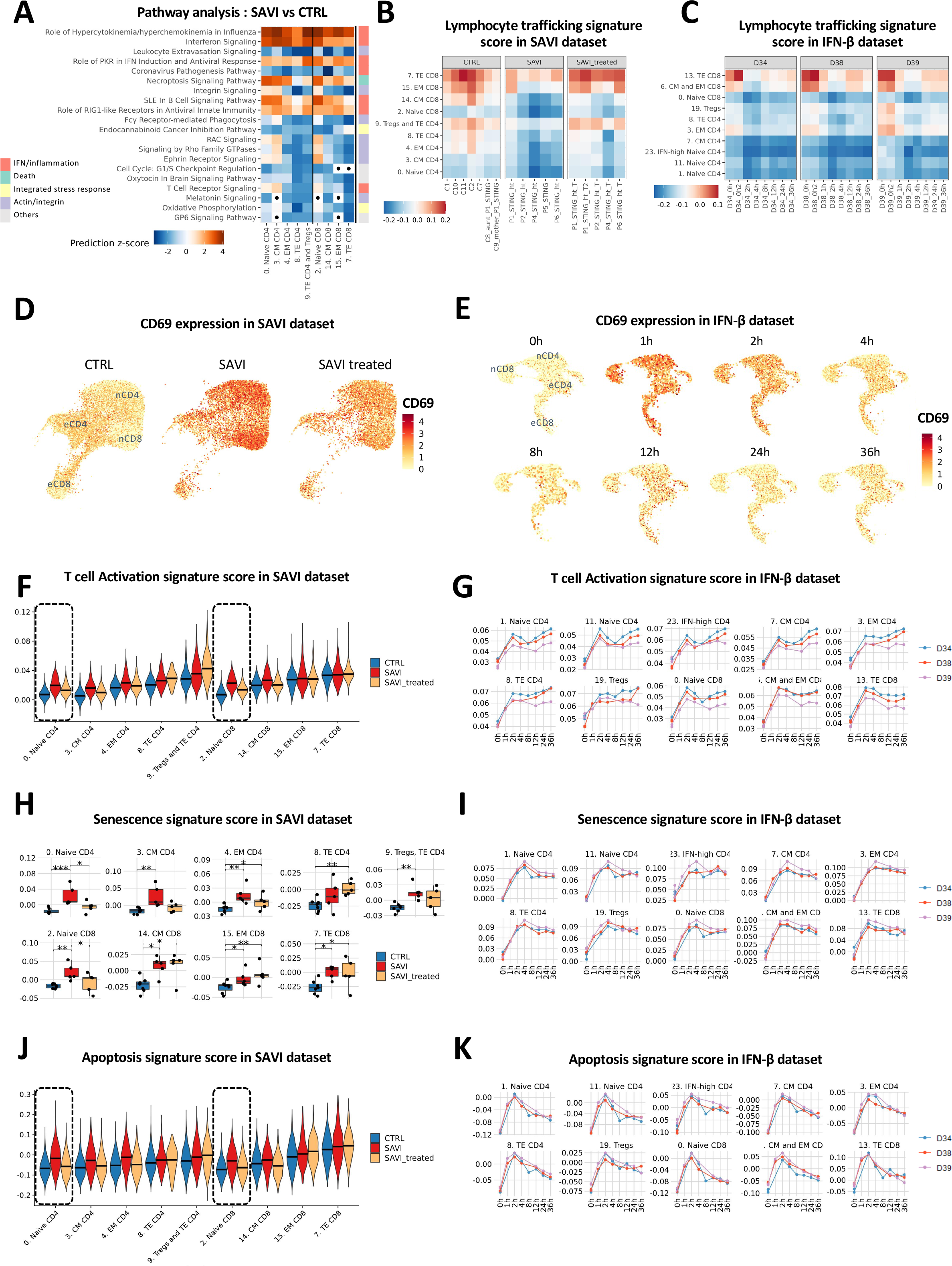
T cells of SAVI have impaired signature score of trafficking machinery, and naive cells have increased activation markers and senescence signature score A. Heatmap of the pathway enrichment analysis performed in IPA, between SAVI and CTRL in each cluster of the CD4+ and CD8+ T cell compartment. Dots indicate non- significant pathways (BH > 0.05). Side color bar indicate groups of pathways based on broader functions. Color scales indicate direction of prediction with orange for predicted activation and blue for predicted inhibition B. Heatmap of a lymphocyte trafficking signature in CD4^+^ and CD8^+^ T cells of the SAVI dataset. Color scales indicate direction of prediction with orange for predicted activation and blue for predicted inhibition C. Heatmap of a lymphocyte trafficking signature in CD4^+^ and CD8^+^ T cells of the IFN-β dataset. Color scales indicate direction of prediction with orange for predicted activation and blue for predicted inhibition D. Feature plot of CD69 mRNA expression level in CD4^+^ and CD8^+^ T cells in the SAVI dataset. nCD4: naïve CD4; eCD4: effector CD4; nCD8: naïve CD8; eCD8: effector CD8 E. Feature plot of CD69 mRNA expression level in CD4^+^ and CD8^+^ T cells in the IFN-β dataset. nCD4: naïve CD4; eCD4: effector CD4; nCD8: naïve CD8; eCD8: effector CD8 F. Violin plot of a T cell activation signature of 4919 genes, in CD4^+^ and CD8^+^ T cells of the SAVI dataset. Dark lines indicate medians G. Evolution of a T cell activation signature of 4919 genes in CD4^+^ and CD8^+^ T cells of the IFN-β dataset over the time course of IFN-β stimulation. Each dot is the average score of the signature for a sample H. Boxplot of a senescence signature of 50 genes, in CD4^+^ and CD8^+^ T cells in the SAVI dataset. Each dot is the average score of the signature for a sample. I. Evolution of a senescence signature of 50 genes in each cluster of the IFN-β dataset over the time course of IFN-β stimulation. Each dot is the average score of the signature for a sample J. Violin plot of an apoptosis signature of 67 genes, in the SAVI dataset. Dark lines indicate medians K. Evolution of an apoptosis signature score of 67 genes in each cluster of the IFN-β dataset over the time course of IFN-β stimulation. Each dot is the average score of the signature for a sample

As T cells of *Sting^N^*^153^*^S/+^* mice were shown to undergo spontaneous activation^11^, we next examined the expression of the early activation marker CD69. T cells of patients displayed a strong increase in the mRNA level of *CD69* (Figures 3D, S3A), mostly observed in the clusters defined as naïve CD4^+^ and CD8^+^ versus effector T cells (Figures 1C, S1E). *CD69* expression was also induced by IFN-β challenge at early time points but returned to basal levels between 4 to 12h post stimulation (Figures 3E, S3B). While the early activation marker remained highly expressed in SAVI patients’ cells and was rapidly downregulated in IFN-β‒stimulated cells, a global T cell activation signature score was increased in both SAVI patients, and IFN-β‒ challenged cells (Figures 3F-G). This activation signature score was most increased in the naïve cells of SAVI patients, while the activation level stayed similar in effector T cells of SAVI patients and CTRLs (Figure 3F). IFN-β stimulates the activation of all clusters of T cells in the IFN-β‒challenged PBMCs (Figure 3G).

The persistent high activation status of naïve T cells associated with a lower proportion of effector cells in SAVI prompted us to hypothesize that naïve T cells failed to expand and differentiate into effector cell upon activation in SAVI and may die. We evaluated a senescence signature as well as the proportion of cells in the Growth and Mitotic phases (G2/M) of the cell cycle (Figures 3H-I, S3C-D). T cells of SAVI patients had an increased senescence signature score compared to CTRLs, which was reduced by JAK-inhibitor treatment in naïve T cells (Figure 3H). A significantly decreased proportion of naïve T cells from SAVI patients in the G2/M phase, partially rescued by the treatment, was also observed, suggesting reduced proliferation capacity compared to CTRLs (Figure S3C), consistent with the impaired T cell proliferation in response to antigens seen in patients. Similarly, in the IFN-β dataset, IFN-β induced increased senescence and decreased proportion of cells in G2/M phase, even though the proportion of cell in the G2/M phase is back to normal after 36h (Figures 3I, S3D). After showing increased activation and senescence, we evaluated if the cells were apoptotic, and revealed increased transcriptomic signature score of apoptosis, particularly in naïve CD4^+^ and CD8^+^ T cells in SAVI patients, restored by JAK-inhibitor treatment (Figure 3J). The IFN-β‒ treated cells showed a sharp increase in apoptosis signature score after 2h of treatment, while the signature score is decreased afterwards (Figure 3K).

Taken together, these data suggest that T cells, identified as naïve based on a set of well-known typical markers, are highly activated in SAVI but have reduced ability to proliferate and differentiate into effector cells, ultimately leading to cell death.

### Identification of disease-associated monocytes in SAVI patients, presenting hyperinflammation and transcriptomic signature of integrated stress response

As the type I IFN response and the NF-κB signaling signature scores were predominantly detected in monocytes and dendritic cells, we next sought to examine these populations in greater details. In addition to the decreased percentages of effector T and NK cells (Figure 1G), we observed a shift in the distribution of classical monocytes, without changes of global monocytes proportions (Figures 1G, S1G, S1I, Figures 4A-B and S4A). One cluster, cluster 17, named “disease-associated cluster”, is composed at 85% of cells from patients, with 66.7% of cells from patients before JAK-inhibitor treatment, and 18.3% of cells belonging to samples obtained from patients under treatment (Figures 4B, S4A). In this disease-associated monocytic cluster (Figure S1E), we observed that both the type I IFN response score and the NF-κB activation score were elevated compared to the classical monocytes (Cluster 5), and that non- classical monocytes (Cluster 12) from SAVI patients also had an elevated score for both signatures, although to a lesser extent (Figure 4C). Expression of type I and type III IFNs were all increased in SAVI patients compared to CTRLs and partially decreased upon JAK-inhibitor treatment (Figure S4B). In SAVI patients, we observed that disease-associated monocytes were the main source of IFN-β mRNA, which was also expressed, but to a lesser extent, by classical and non-classical monocytes (Figures 4D, S4C). IFN-α mRNA subtypes were also expressed by disease-associated, classical and non-classical monocytes, as well as pDCs. To better understand the molecular mechanisms underlying the disease-associated cluster, we performed differential expression analysis between the disease-associated monocytes of SAVI patients (cluster 17) and their classical monocytes counterparts (cluster 5). We observed 984 DEGs (Figures 4E, S4D, supplementary file 5), with the upregulation of key players involved in the integrated stress response (ISR), cell activation and transcripts of proinflammatory cytokines including *IL1B* and transcripts of several type I IFN (such as *IFNB1*, *IFNA1*, and *IFNA2*). Pathway analysis revealed a strong enrichment of genes negatively regulating the EIF2 signaling pathway, which can be associated to an increase of the ISR^24^ (Figures 4F, S4D, supplementary file 6). To validate the upregulation of the ISR, we looked at the mRNA level of *PPP1R15A*, which encodes the ISR marker GADD34. *PPP1R15A* expression reached the highest level in the disease-associated monocytes of the SAVI group (Figures 4E and G). An unfolded protein response (UPR) signature, known to contribute to the ISR, obtained from *Raich* et al^25^, followed a similar pattern with a high score in patients’ disease-associated monocytes. Of note, non-classical monocytes of patients expressed increased *PPP1R15A* or UPR signature score. JAK-inhibitor treatments seem to partially reduce these pathways in both non-classical and disease-associated clusters (Figure 4G). In the stimulated monocytes of the IFN-β dataset both *PPP1R15A* and the UPR signature score were also increased after 36h of stimulation (Figure S4E). Among the other pathways upregulated in this disease-associated cluster, we also report pathways linked to response to IFN, production of inflammatory cytokines, and cell death along with an enrichment of genes involved in oxidative phosphorylation (Figure S4D). Overall, our data suggest that this disease-associated monocytic cluster corresponds to classical monocytes that display a high level of inflammation and ISR, at the transcriptomic level and likely related to the constitutive STING activation.

**Figure 4:**
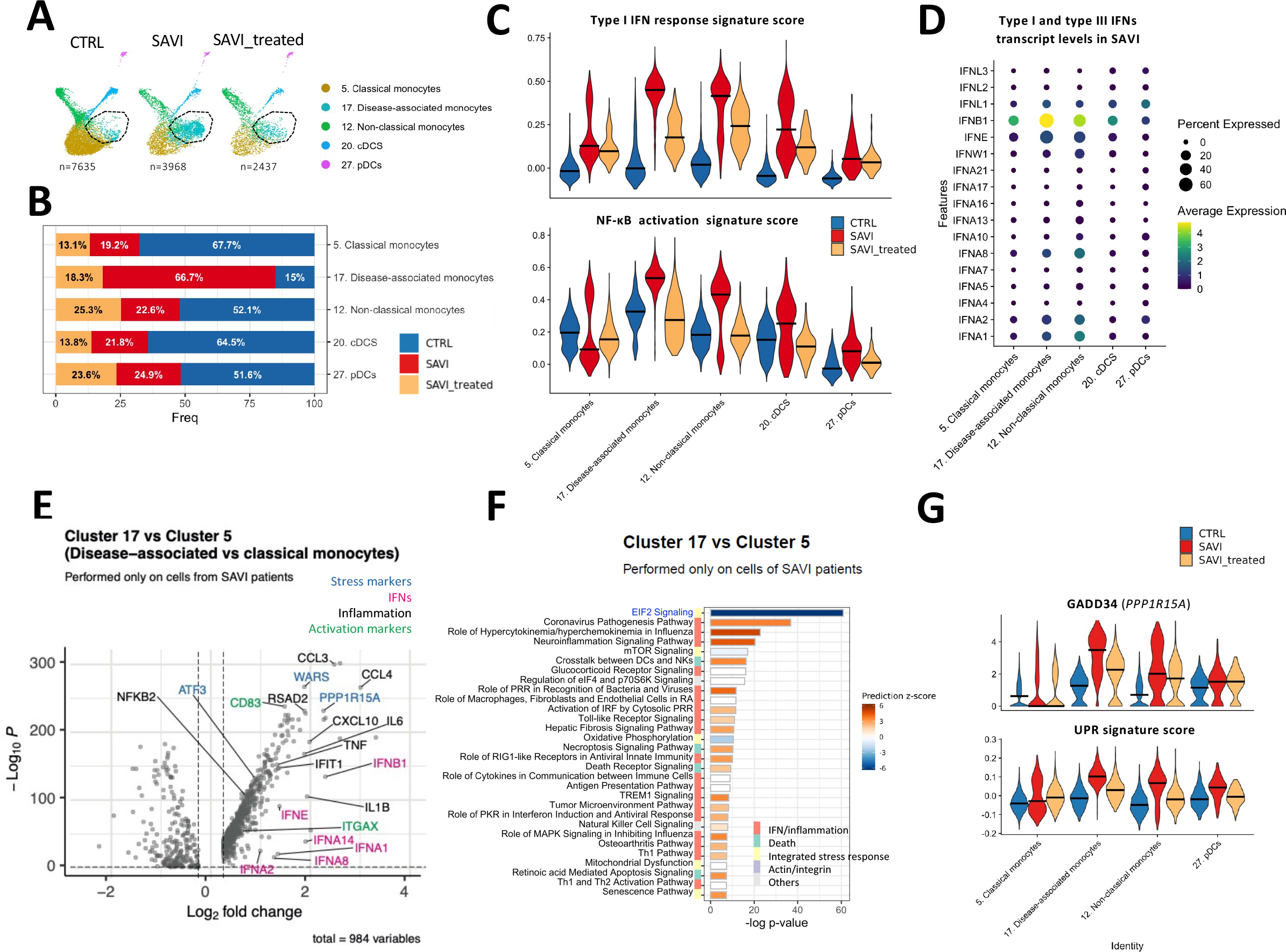
*In silico* clustering of monocytes and DC populations reveals a disease-associated cluster of monocytes characterized by elevated signature score of type I IFN response, NF-κB activation and integrated stress response A. UMAP of the monocytes and dendritic cells clusters in the SAVI dataset, separated by groups (CTRL, SAVI, or SAVI_treated). Black circle indicates a cluster of cells enriched in the patients termed “disease-associated monocytes.” B. Composition of monocyte and dendritic cell clusters in each group of the SAVI dataset (CTRL, SAVI, or SAVI_treated) C. Violin plots of the score of a type I IFN response signature of 272 genes (top) and of an NF-κB activation signature of 200 genes (bottom) in the SAVI dataset. Dark lines indicate medians D. Dotplot of the expression levels of all type I and type III IFNs, in each monocyte and dendritic cell clusters, in the SAVI group. E. Volcano plot of the differentially expressed genes between the cells from SAVI patients in cluster 17 (disease-associated monocytes) and cluster 5 (classical monocytes). Stress, IFN and activation related genes are respectively highlighted in blue, pink and green. F. Pathway enrichment analysis, performed in IPA, between the cells from SAVI patients in cluster 17 (disease-associated monocytes) and cluster 5 (classical monocytes). Side color bar indicate groups of pathways based on broader functions. Color scales indicate direction of prediction with orange for predicted activation and blue for predicted inhibition G. Violin plot of the expression of *PPP1R15A* which codes for GADD34 (top) and an unfolded protein response signature of 85 genes (bottom), in the SAVI dataset. Dark lines indicate medians

### Inference of cell-to-cell communication predicts that hyperinflammatory monocytes in SAVI could drive hyperactivation and death of effector T cells

To better understand how monocytes from SAVI patients may influence T cells (Figures 3 and 4), we sought to infer monocytes-to-T cell communications. To this end, we used the ICELLNET framework^26^, which infers a score of cell-to-cell communication based on the transcriptomic level of ligands expressed by a sender cell type (here, both clusters of classical monocytes; cluster 5. Classical monocytes, and cluster 17. Disease-associated monocytes), and the transcriptomic level of their receptors expressed by a receiving cell type (here, T cells; Figure 5A). We applied this framework to the SAVI dataset to infer the communication between classical monocytes (including the disease-associated cluster) and each CD4^+^ and CD8^+^ T cell cluster in the SAVI group and the CTRL group separately (Figure 5B). We reproduce a similar analysis on the IFN-β dataset for each time point (Figure 5C), which allowed us to decipher IFN-dependent and IFN-independent predicted interactions, once compared to the SAVI dataset. Hierarchical clustering of the T cells clusters based on their communication score with monocytes showed a clear separation between clusters from untreated SAVI patients and healthy donors (Figure 5D). We observed a predicted increase of type I IFN-IFNAR interactions in SAVI patients, which most probably reflects the drastic upregulation of IFN transcripts in the monocytes of SAVI patients (Figure 4D), although this interaction is not predicted in the IFN-β dataset (Figures 5D, 5E). Other chemokines and proinflammatory cytokines including CXCL9, CXCL10, and CXCL11 with their common receptor CXCR3, as well as the pair IL-6/IL-6R, are predicted to have increased interactions in a type I IFN-dependent manner (Figures 5D, 5E). In contrast, the increase of IL-12 and IL- 27 interactions with their receptors is likely independent of type I IFNs, as absent from the predictions in the IFN-β dataset.

**Figure 5:**
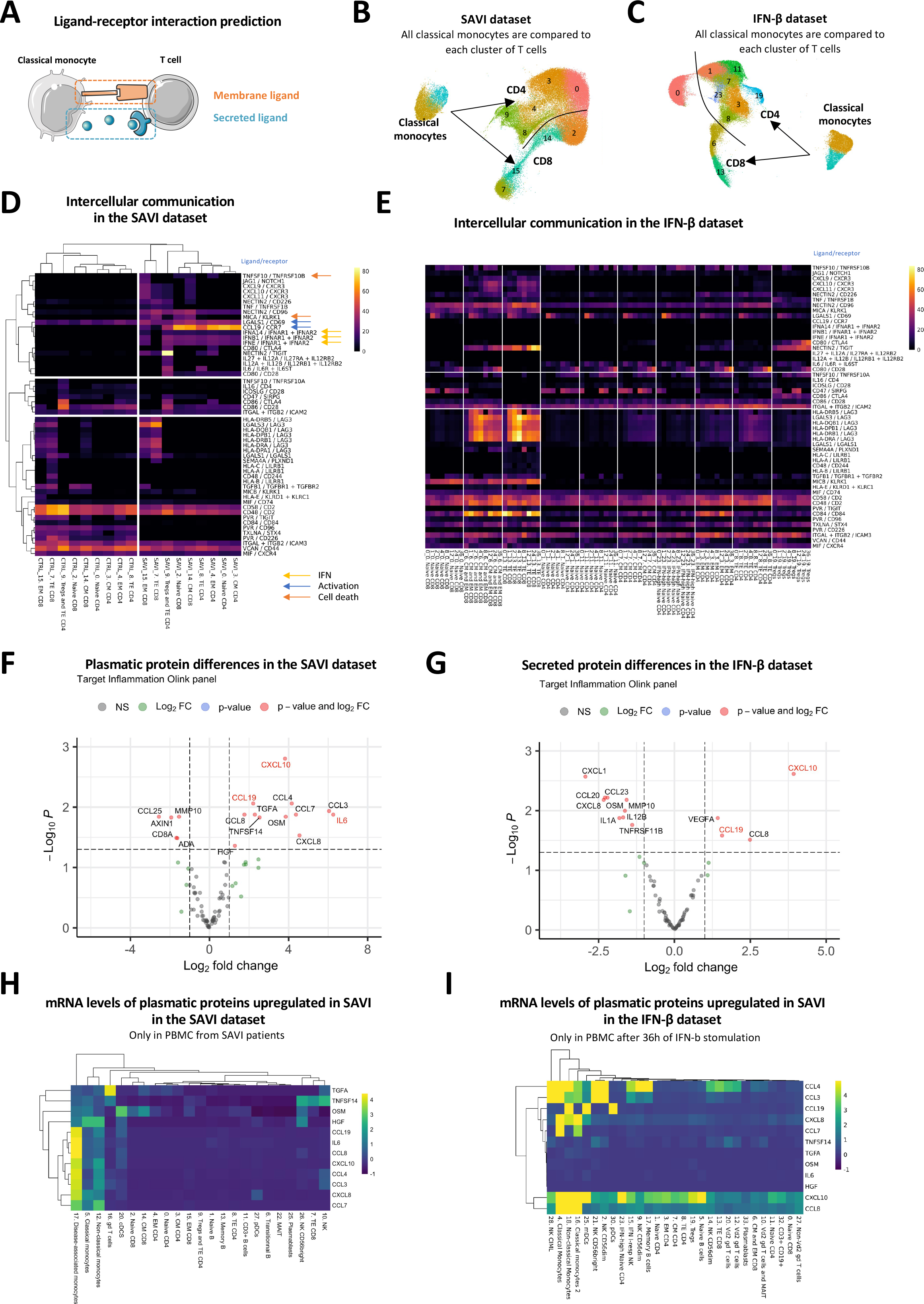
Inference of cell-to-cell communication based on scRNA-seq datasets suggests that hyperinflammatory monocytes of SAVI drive activation and death of T cells through cytokine secretion A. Cell-to-cell communications are inferred from the expression of ligands (secreted or presented at the membrane) expressed by a sender cell (here monocytes) and receptors expressed by a receiver cell (here, T cells) B. In the SAVI dataset, we evaluate the interactions between both cluster of monocytes (cluster 5 and cluster 17) and each cluster of the T cell compartment C. In the IFN-β dataset, we evaluate the interactions between both cluster of monocytes (cluster 4 and cluster 16) and each cluster of the T cell compartment D. Heatmap of the score of each ligand/receptor pair between each T cell cluster and the monocytes, either in the SAVI or the CTRL group. Hierarchical clustering based on Pearson correlation. Colored arrow indicate association to specific pathways. E. Heatmap of the score of the same ligand/receptor pair as observed in the SAVI dataset, between each T cell cluster and the monocytes, in each timepoint of the IFN-β dataset F. Volcano plot of the differential protein secretion between SAVI patients and associated CTRL for 96 proteins measured from the plasma. Genes written in red are in agreement with cell-to-cell interaction predictions. G. Volcano plot of the differential protein secretion between PBMCs stimulated with IFN-β for 36h and unstimulated for 96 proteins measured from the plasma using the Olink Inflammation panel. Genes written in red are also increased in the plasma of SAVI. H. Heatmap of the scaled mRNA levels of the 12 proteins found upregulated in the blood of SAVI patients in panel F, in each cell type in the SAVI patients I. Heatmap of the scaled mRNA levels of the 12 proteins found upregulated in the blood of SAVI patients in panel F, in each cell type in cell treated with IFN-β for 36h

Strikingly, the strongest predicted interaction in SAVI patients was reported for the CCL19/CCR7 ligand/receptor couple, which plays an essential role in T cell activation^27^. This communication was not predicted to occur in healthy PBMCs treated with IFN-β, nor in SAVI patients under JAK-inhibitor treatment (Figures 5E, S5A), arguing for a mechanism independent of type I IFN production. Checkpoint interactions, such as CD80/CD28, Nectin2/CD226, Nectin2/TIGIT, and Nectin2/CD96, involved in T cells stimulations^28^, were also increased in both SAVI patients and following IFN-β stimulation, but should be interpreted with caution as monocytes are not professional antigen presenting cells. Moreover, our analyses predicted enhanced crosstalk between LAG3, involved in immune exhaustion and regulation of effector function^29^, and its ligands LGALS3 and MHC class II molecules in SAVI, corresponding to the strongest induction predicted in the IFN-β dataset (Figure S5B). We also observed in SAVI patients and IFN-β stimulated cells increased communications involved in apoptosis induction with TNFSF10/TNFRSF10B and LGALS1/CD69 (Figures 5D, 5E)^30, 31^. In CD8^+^ T cell interactions with monocytes, the pair MICA/KLRK1 was predicted to be increased in SAVI patients. MICA is a stress-induced ligand belonging to the MHC-I family and known to lead to cell lysis when bound to its receptor KLRK1/NKG2D^32^ (Figure 5D). The same interaction is also predicted to be strongly induced by IFN-β‒stimulation exclusively in CD8^+^ T lymphocytes. Overall, these data point towards a role of the disease-associated monocytes in favoring T cells chronic activation and death.

### Circulating cytokines in the plasma of SAVI patients as biomarkers of the disease

As several cytokines/chemokines were seen upregulated in SAVI patients at the transcriptomic level, we evaluated the level of 96 inflammation-related proteins in the plasma of SAVI patients and associated CTRLs using the proteomic proximity extension assays (PEA). We confirmed the previously known increase of IL-6^16^, and we observed increased plasmatic levels of CXCL10 and CCL19 (Figure 5F), which were predicted to play a role in monocytes-to-T cell communications (Figure 5D). Other proinflammatory cytokines, such as CCL3, CCL4, CCL7, and CCL8, were also increased in the plasma of SAVI patients compared to CTRLs (Figure 5F). The secretory profile of PBMCs stimulated with IFN-β for 36h was characterized by induction of CXCL10, VEGFA and CCL19 (Figure 5G). Although IFN-β‒stimulation increased secretion of CXCL8, we observed a decrease in SAVI patients. To assess the correspondence between the protein and the mRNA levels of the cytokines and chemokines measured in the plasma of SAVI patients, we evaluated the expression of the 12 cytokines found to be increased in the plasma of SAVI patients in the scRNA-seq data (Figure 5H). Our results suggest a strong role of the disease-associated monocytes in the secretion of several of these cytokines and therefore validate our cell-to-cell inference predictions, even though the transcripts levels of some of these cytokines were close to the limits of detection (Figures 5H, S5C). Of note, *CCL3*, *CCL4* and *IL-6* were among the most upregulated genes in the disease-associated monocytes (Figure 4E). PBMCs stimulated with IFN-β had a more homogeneous expression of inflammatory cytokines across cell types, in particular monocytes, DCs, and NK cells (Figures 5I, S5D). The specific elevation of type I IFN-independent cytokines, in particular CCL3, CCL4 and IL-6, suggests that they may be used as blood biomarkers of SAVI to aid the differential diagnosis with other type I interferonopathies, but this will remain to be further explored.

### A transcriptomic signature specific of STING activation and independent of response to IFN-β

Our next objective was to try to identify a transcriptomic signature specific of STING activation, but independent of response to IFN-β stimulation. The workflow used to obtain such a signature is described in Figure 6A, and supplementary file 7. As our *in-silico* clustering analysis revealed a group of monocytes specific to the disease with elevated inflammation and ISR (Figure 4), we aimed to establish a transcriptomic signature based on the genes driving the differential clustering of these cells. The first step therefore consisted in the extraction of the genes specific to the disease-associated cluster (Step 1, Figure 6B). We obtained 95 genes that are highly upregulated in the disease-associated cluster compared to the other cluster of classical monocytes in SAVI patients (Cluster 17 vs Cluster 5, only on SAVI cells, FDR <0.05, log2FC > 1). One gene, *G0S2*, was filtered out from the list as its expression was not increased in SAVI compared to CTRLs (Step 2, Figure 6C). To obtain a signature independent of type I IFN response, an important step of the gene selection was to remove all genes related to type I IFN and type I IFN response. We started by removing all type I IFN transcripts that were found in the list (namely *IFNA1*, *IFNA8*, *IFNA14*, *IFNB1*, and *IFNE*) (Step 3, Figure 6C) and then ISGs. To this end, we identified the genes upregulated by IFN-β in each timepoint of the IFN- β dataset. We removed 68 genes that were increased by type I IFN in monocytes and DCs in at least one of the timepoints of the IFN-β dataset (Step 4, Figure 6D). Consequently, 21 genes remained in the STING activation signature (Figure S6A). These genes were mostly associated with TNF-α signaling via NF-κB (Figure 6E). We confirmed that this signature was specific to the SAVI patients, as compared to our CTRL group, partially reduced in JAK-inhibitor-treated patients (Figure 6F). We also confirmed in the IFN-β dataset that the resulting signature score is not increased by type I IFN at any timepoint (Figure S6B).

**Figure 6:**
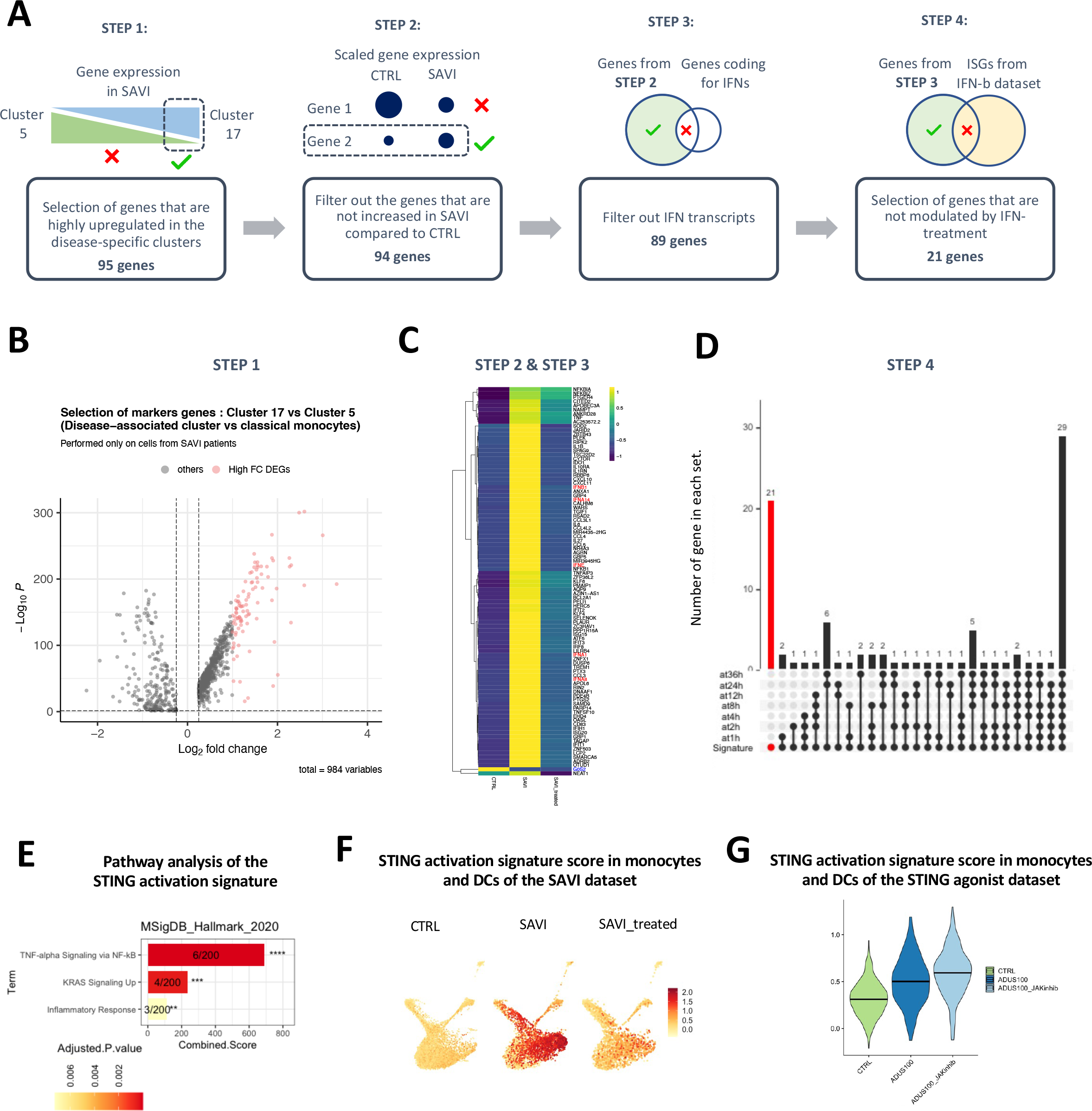
Design of a STING-activation signature, independent of type I IFN response, based on the transcriptome of the disease-associated monocytes A. Workflow used for the extraction of an IFN-independent STING-activation signature. B. Volcano plot of the DEGs between cluster 17 and cluster 5, in SAVI. The 95 genes highlighted in pink have log2FC > 1 and were selected as the basis of the STING activation signature C. Heatmap of the 95 genes of selected in panel B, in the monocyte and DCs of CTRL, SAVI and SAVI treated. Blue writing represents the genes whose expression is higher in CTRL than SAVI and are filtered out in Step 2. Red writing represents the IFN transcripts that are filtered out in Step 3. D. Upset plot of the genes of the STING-activation signature. This graph represents set intersections by depicting how many of the 89 genes of the STING activated signature are also found in the lists of genes upregulated as a response to IFN-β. Dots in the bottom part display which interaction is represented and bars in the upper part show how many genes are found in this interaction. The red bar represents the genes that are uniquely found in the signature and not upregulated by IFN-β at any timepoint. These 21 genes are kept in the final STING activation signature, while others are removed in Step 4. E. Bar charts of the pathways predicted in EnrichR to be significantly enriched for the 21 genes of the STING activation signature in MSigDB_Hallmark_2020 database. Pathways are ranked based on combined score; *(p < 0.05), **(p < 0.01), ***(p < 0.001), **** (p < 0.0001) F. Feature plot of the signature score of the 21 genes of the STING activation signature in the monocytes and DCs of CTRL, SAVI and SAVI_treated in the SAVI dataset G. Violin plot of the signature score of the 21 genes of the STING activation signature in the monocytes and DCs of CTRL, ADUS100 and ADUS100+JAK-inhibitor in the ADUS100 dataset. Dark lines indicate medians

To confirm the relevance of this signature in the context of STING activation, and to evaluate whether the signature is indeed type I IFN-independent, we designed an *in vitro* model of STING activation, in which we used PBMCs from a healthy donor, either unstimulated, stimulated with ADUS100 (a STING agonist), or with a combination of ADUS100 and JAK-inhibitor (inhibitor of type I IFN signaling). After performing scRNA-seq, we evaluated STING activation by measuring the score of type I IFN response and NF-κB activation, which were both increased by ADUS100 and dampened by the addition of a JAK-inhibitor (Figure S6C). We observed a strong increase of the STING activation signature score in monocytes and DCs following ADUS100 stimulation, which remained elevated when blocking the response to type I IFN (ADUS100 + JAK-inhibitor) (Figure 6G). The STING activation signature score was restricted to monocyte and DC populations in both the SAVI patients (Figure S6D) and the ADUS100 stimulated PBMCs (Figure S6E).

Overall, we propose that this list of 21 genes may represent a signature specific of STING activation and independent of response to type I IFN.

### Identification of a second genetic variant in PERK in a SAVI patient with low UPR

While analyzing the transcriptomic profiles of the SAVI patients, we identified several pathways that were differentially modulated in P1. We noted, in this patient, a marked reduction of the UPR, which was largely increased in other patients (Figure 7A). Of note, IFN- β stimulation induced a slight increase of the UPR (Figure S7A). This observation is accompanied by a very low expression of the ISR marker *PPP1R15A*, which encodes GADD34, in monocytes and DCs (Figure 7B). Further exploration showed differences in induction of the apoptosis signature and NF-κB signaling, though less drastic than the UPR (Figure 7C, 7D). These differences are even observable when compared to P2, a relative of P1 (Figure S7E). In contrast, the type I IFN response is found similarly increased in P1 as compared to other SAVI patients (Figure S7B, S7C, S7D).Of note, P1 is the only patient in this study to present with a systemic lupus erythematosus (SLE)-like phenotype^1^, in addition to its SAVI phenotype.

**Figure 7:**
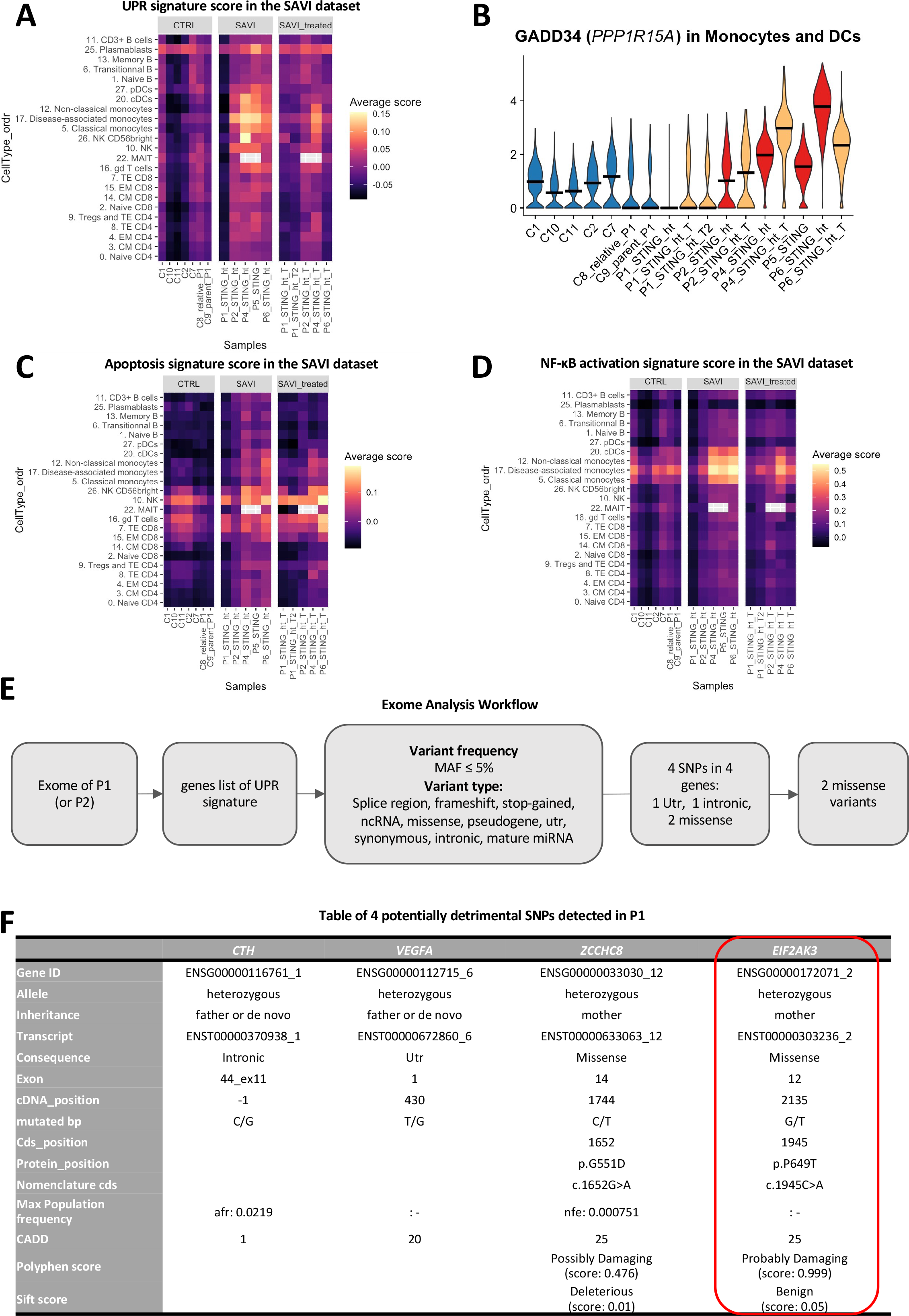
Exome analysis of a patient with low UPR reveals another SNP in *EIF2AK3* A. Heatmap of a UPR signature of 85 genes in each sample and each cluster of the SAVI dataset. Grey squares are used when no cells from a sample in found in a cluster. B. Violin plot of the expression of *PPP1R15A* which codes for GADD34 in monocytes and DCs of the SAVI dataset. Dark lines indicate medians C. Heatmap of an apoptosis signature of 67 genes in each sample and each cluster of the SAVI dataset. Grey squares are used when no cells from a sample in found in a cluster. Grey squares are used when no cells from a sample in found in a cluster. D. Heatmap of an NF-κB activation signature of 200 genes in each sample and each cluster of the SAVI dataset. Grey squares are used when no cells from a sample in found in a cluster. Grey squares are used when no cells from a sample in found in a cluster. E. Workflow of the exome analysis based on the 85 genes of the UPR signature. MAF: Major Allele Frequency. F. Table of the four SNPs found in the patient with low UPR

The drastic default of UPR induction in P1 compared to the other patients, even to the relative P2, prompted us to reanalyze the exome sequencing previously performed for this patient, including parents and P2^1^. A supervised analysis was performed using a list including the 85 genes of the UPR signature (Figure 7E). Exome analysis revealed single-nucleotide polymorphisms (SNPs) in four genes: *CTH*, *VEGFA*, *ZCCHC8* and *EIF2AK3* (Figure 7F). Variants in *CTH* and *VEGFA* are intronic or in untranslated regions and therefore, may not induce a change in protein function. In contrast, variants in *ZCCHC8* and *EIF2AK3* induced core changes in amino acid chain and may be of importance. While the STING p.V155M mutation was inherited the affected parent, both UPR genes variants were inherited from the healthy parent suggesting that those mutations could play a role in the transcriptomic differences observed between this patient and the relative P2, regarding the UPR signature score (Figure S7E). The first SNP in *ZCCHC8* was a missense variant (p.G551D). Interestingly, the protein encoded by this gene was previously reported to interact with STING in mouse bone marrow-derived DCs ^33^. Moreover, an autosomal dominant *ZCCHC8* mutation was reported in patients with pulmonary fibrosis^34^, a clinical feature observed in P1. However, this variant had an allele count in the population higher than expected for a pathogenic variant (it was seen 110 times in gnomAD giving an allele frequency of 0.000405). Of note, advanced modeling of protein sequence and biophysical properties indicated that this missense variant is not expected to disrupt ZCCHC8 protein function (Clinvar 1357971). Finally, ZCCHC8 appeared to be a downstream target of the UPR pathway. Therefore, this variant was no longer considered as a possible cause of the overall reduced UPR signature score observed in P1 (Figure 7A).

The last SNP was a private missense variant (p.P649T) in *EIF2AK3*, which encodes PERK, a key activator of the UPR. This variant was confirmed by Sanger sequencing (Figure S7F). This protein was recently shown to be activated upon STING activation through direct interaction between the C-terminal tail of STING and the kinase domain of PERK^20^. Once activated, PERK induces the phosphorylation of key components of the ISR. The p.P649T variant carried by P1 is in the kinase domain of PERK, which carries the ISR-inducing function. This variant induces a change in residue type from a nonpolar amino acid (Proline) to a polar amino acid (Threonine) and all prediction scores suggested that this variant is probably damaging. Importantly, the UPR signature score of the parent who carries the variant (C9) appeared in T and B lymphocytes slightly lower than the score observed in the healthy relative (C8) who is not carrying the *EIF2AK3*variant (Figure 7A). Overall, these findings were supporting a potential causal effect of the p.P649T variant of PERK in the reduction of the UPR signature in P1.

## Discussion

In the present study, we combined single-cell high-throughput analysis of PBMCs from five SAVI patients before and after initiation of JAK-inhibitor treatments, with the analysis of *in vitro* IFN-β-stimulated PBMCs from healthy donors (1h to 36h). We highlighted the main pathways dysregulated in the immune cells of SAVI patients, including type I IFN and non-type I IFN-mediated inflammation, an exacerbated integrated stress response and cell death. We revealed a persistent T cell activation, and confirmed a previously reported decreased proportion of circulating effector T cells^1, 14^. Furthermore, we identified, through *in silico* clustering analyses a group of disease-associated monocytes, characterized by elevated integrated stress response and high inflammation. Cell-to-cell communication inferences based on our single-cell datasets predicted a potential role for these highly inflamed monocytes in the observed phenotypes of T cells in the blood of SAVI patients. We extracted from the disease-associated monocytes a list of 21 genes specific to the activation of STING but independent from the type I IFN response. We also reported in the plasma of SAVI patients a specific elevation of CCL3, CCL4 and IL-6 independent of a primary stimulation by type I IFN. Together, our findings provide a deeper understanding of the SAVI pathogenesis.

T cell lymphopenia is a clinical hallmark of SAVI^1, 14^. We have confirmed the drastic reduction of the frequency of effector T cells in our cohort of SAVI patients. By exploring, at the single cell level, other T cells subsets found in the blood of SAVI patients we highlighted a unique phenotype in clusters defined by transcriptomic markers as naïve T cells. We revealed that these naïve T cells seemed to be maintained in an early activation state as measured by CD69 expression. This early T cell activation is observed as early as 1h after IFN-β stimulation, but strongly downregulated after 4h. These results are in agreement with previous reporting suggesting that T cells carrying a gain-of-function *STING1* mutation may be constitutively activated as a result of homeostatic reactivity to self-antigen^11, 35^. Additionally, we could hypothesize that T cells are activated in an antigen-independent manner by inflammatory cytokines including type I IFN^36^. While activation should prompt T cell proliferation, we report increased activation of the senescence pathways in naïve T cells of SAVI patients, associated with a decrease in the proportion of cells reaching the G2M phase of the cell cycle. Several studies have highlighted the ability of chronic IFN-β to trigger senescence^37, 38^. Our observations from the IFN-β dataset support these studies, although the decrease in cell cycle is only transiently observed. These results concur with the antiproliferative role of STING^14^, as well as its pro-senescence^10, 39^ and pro-apoptotic^12^ roles. Altogether, our results could suggest that the decrease of effector T cells may result from the altered naïve T cell compartment. We propose that naïve T cells could lose their ability to transition into an effector state due to an hyperactivation state combined with premature senescence and apoptosis.

*In silico* clustering analysis of the SAVI dataset revealed a disease-associated cluster composed of 85% of cells from SAVI patients, mostly before treatment, suggesting that these cells might be clustering together due to genes and pathways dysregulated by STING activation. This cluster was of classical monocytes origin, and presents an increased expression of activation and inflammatory markers. These disease-associated monocytes had elevated *GADD34* gene expression, an increased UPR signature score, and decreased ribosomal genes expression, which altogether imply the activation of the ISR. While our results did not allow us to conclude on the exact mechanism driving the increase of ISR, the recent discovery of the direct activation by STING of an ISR mediator, PERK, suggests that this could be mediated through PERK^20^. PERK is an ER-stress sensor that, like inactivated STING, is found on the ER membrane. Upon unfolded protein sensing, PERK phosphorylates eIF2α, thereby triggering the ISR. Its direct activation by STING, independently of unfolded proteins, has recently been demonstrated in human cell lines and in mice^20^. These observations suggest that elevated UPR in this cluster reflects a STING-induced activation of PERK. However, as PERK is not the only trigger for UPR, we cannot exclude that other mechanisms are involved in high UPR in these disease-associated monocytes.

Intriguingly, we observed in the patient P1, as opposed to other patients, a low UPR signature score. This SAVI patient is the only one in our cohort who had lupus-like manifestations in addition to a more typical SAVI phenotype (skin vasculopathy and lung inflammation). Exome analysis of the 85 genes of the UPR signature revealed a missense p.P649T variant in *EIF2AK3,* encoding PERK, in this patient, inherited from the healthy parent. Prolines are important residues for protein backbone^40^, often found in tight turns in protein structures. The P649 residue is found between an α-helix and a β-sheet in the cytoplasmic kinase domain. Substitution with a threonine may change the structure of the protein and affect the kinase activity. This residue lies in the kinase domain of PERK, which is responsible for eIF2α phosphorylation, a key event for the initiation of the ISR. This variant was predicted to be highly deleterious. In addition, this variant had never been described, suggesting that this private variant might be highly detrimental in combination with STING-activating mutations.

We found some cells in the P1 patient with high UPR and identified these as plasmablasts. Interestingly, it had been shown that plasmablasts were characterized by high UPR due to their essential secretory role, but in a PERK-independent manner^41, 42^. The fact that plasmablasts of the PERK-mutated patient retained their ability to activate the UPR, further points to a PERK-dependent mechanism for the activation of the UPR in disease-associated monocytes. We concluded that the unique UPR defect in the P1 patient may be associated with the SNP located in the PERK kinase domain. To go further, the kinase domain of PERK is responsible for eIF2α phosphorylation, a key event for the initiation of the ISR which overall strengthen the importance of PERK mediated ISR in disease-associated monocytes of SAVI patients.

In many autoinflammatory diseases, the cellular source of type I IFN is still debated. While it could be assumed that pDCs, which are professional IFN-producers, are the main source of type I IFN in inflammatory diseases, they have also been suggested to gradually lose their ability to produce IFN-α in lupus-prone mice^43, 44^. Here, we report that several cell populations express transcripts of type I IFNs in SAVI patients: classical monocytes, non-classical monocytes, and disease-associated monocytes. These results partially corroborate the previous finding that pDCs and monocytes expressed IFN-α proteins in SAVI patients^45^. Our data extend this knowledge by indicating that disease-associated monocytes show the highest expression of IFN-β at the mRNA level.

By leveraging an expert-curated database of intercellular communications to infer monocytes-to-T cells interactions^26^, we provided clues that hyperinflammatory monocytes could promote activation of T cells, as well as cell death. This method revealed the establishment of T cell-activating signaling in SAVI patients. Indeed, we note the increase of CCL19/CCR7 and LGALS1/CD69, which play a role in T cell activation^27, 30^. Several of the communications inferred involved checkpoint interactions. While monocytes are not antigen-presenting cells (APCs) themselves, recent work has suggested the ability of activated monocytes to activate CD8^+^ T cells^46^. Additionally, they are the precursor of monocyte-derived macrophages and dendritic cells, which are APCs^47^. The increase of transcripts of antigen-presentation in blood monocytes may reflect the enhanced capacity of these monocytes-derived presenting cells to effectively activate T cells. Additionally, we note several communications that trigger T cell death, such as TRAIL(*TNFSF10*)/TNFRSF10B^48^ or MIKA/KLRK1^32^, or involved in immune exhaustion such as LAG3 and its ligands^29^. These data point towards a potent role of the monocytes to prime T cells towards activation and cell death. Although these interactions were predicted at the transcriptomic level, several of the cytokines involved in the predicted monocytes-to-T cells communications were also elevated at the protein level in the plasma of SAVI patients. We have also shown that most of the cytokines that we have found increased were likely secreted by the disease-associated monocytes, by evaluating their expression at the mRNA level in both the SAVI and the IFN-β single-cell datasets.

Further studies will be necessary to investigate the contribution of other cell types that were not included in this work, such as neutrophils or epithelial and endothelial cells, which may also play a role in the secretory profiles of the blood of SAVI patients^49^. Nonetheless, our results converge towards a central role of these disease-associated monocytes in SAVI, particularly in the priming of T cells towards hyperactivation and apoptosis.

To further evaluate the relevance of the disease-associated cells in the pathogenesis of SAVI, we suggest that their presence should be investigated in the lungs of SAVI patients. Indeed, monocytic infiltrate has been observed, and it is conceivable that they could be composed of the disease-associated monocytes identified in the blood. The role of monocytes and monocyte-derived macrophages is increasingly recognized in pulmonary fibrosis^50–53^. Additionally, the integrated stress response, a strongly induced pathway in these monocytes, has also been proposed as an important driver of pulmonary fibrosis^54^, which further supports the hypothesis that the disease-associated monocytes may drive the pulmonary damage in SAVI patients. If these disease-associated monocytes could be indeed found in the lungs of SAVI patients, we propose that they may constitute an interesting indication for treatment monitoring. Other potential targets of interest in SAVI are PERK and the ISR, which are the targets of compounds under development for other diseases^55^.

Four patients were treated with JAK-inhibitors (mainly Ruxolitinib, a JAK-1/2 inhibitor). While JAK-inhibitors are beneficial to SAVI patients, they do not allow for complete remission, and patients with advanced lung disease prior to treatment show little to no amelioration^3^. Here, we have assessed the impact of JAK inhibition on each pathway found to be dysregulated in our SAVI patients prior to treatment and our data support an incomplete impact of the treatment at the molecular level. In T cells, while JAK-inhibitor treatments seemed to correct the senescence and apoptosis observed in naïve T cells, they did not allow to restore the proportion of T cells to normal levels. Notably, Tofacitinib was recently suggested to inhibit T cell proliferation^56^, consistently with JAK3 inhibition, and further work is needed to evaluate if JAK-1/2 inhibitors also induce the same effect.

The current diagnosis of type I interferonopathies, including SAVI, relies on clinical evaluation followed by measurement of the level of expression of IFN-stimulated genes in PBMCs or whole blood of suspected patients. This approach is common to all type I interferonopathies, and subsequent sequencing (either Sanger sequencing or exome sequencing) is required to pinpoint a specific gene^1^. There is therefore an unmet need for an easy-to-use, differential diagnosis strategy between the different type I interferonopathies. We noticed the elevation of CCL3, CCL4 and IL-6 at the transcriptomic level in the disease-associated monocytes and at the protein level in the plasma, and we propose that they be added to the panel of 6 ISGs routinely measured at the transcriptomic level. An elevated cytokine expression of these cytokines associated with high ISG score could lead to a suspicion of *STING1* gain-of-function mutation.

In addition, thanks to the identification of a monocytic derived disease-associated cluster, we were able to identify, at the transcriptomic level, a signature driven by STING activation, independent of type I IFN response. It will be interesting to test if this signature could serve as another tool to discriminate SAVI patients from other interferonopathies such as Aicardi-Goutières Syndrome or COPA patients. Further work is needed to evaluate the relevance of this signature in the differential diagnosis of SAVI against other type I Interferonopathies, but interestingly, *IL1B*, which encodes the inflammasome activation marker IL-1β, is part of the STING activation signature, which is consistent with the previously described role of STING in NLRP3 inflammasome activation^57^. Several genes of the signature are linked to NF-κB activation, reinforcing the association of this signature to type I IFN-independent roles of STING.

In summary, our study represents the first single-cell RNA sequencing analysis performed in the hematopoietic compartment of SAVI patients. We propose that the loss of effector T cells in SAVI patients could be consequence of an altered ability of naïve cells to differentiate and highlight a pathogenic group of monocytic cells characterized by an elevated integrated stress response. Moreover, we identified a signature specific of STING activation and independent of type I IFN response that should be further studied to evaluate if it could aid better distinguish SAVI from other type I interferonopathies. Additionally, we have leveraged scRNA-seq differential analyses between SAVI patients to point towards the discovery of a potential new variant in PERK, which could help to better explain the differences observed at the clinical level in this SAVI patients, a step further toward personalized medicine.

## Data Availability

The scRNA-seq data from the SAVI and the IFN-β datasets have been deposited in GEO and will be publicly available upon paper publication, under the superseries number GSE226601, with respective series number GSE226598 and GSE226572 for the SAVI and IFN-β datasets respectively.

https://www.ncbi.nlm.nih.gov/geo/query/acc.cgi?acc=GSE226601

## Acknowledgements

The study was supported by the Institut National de la Santé et de la Recherche Médicale (INSERM), by the Atip-Avenir, by government grants managed by the Agence National de la Recherche as part of the “Investment for the Future” program (Institut Hospitalo-Universitaire Imagine, grant ANR-10-IAHU-01, Recherche Hospitalo-Universitaire, grant ANR-18-RHUS- 0010), by a CIFRE PhD program with Sanofi, by a Sanofi iAward Europe, and the “Emergence ville de Paris” program. C.C. and L.D are both the recipients of CIFRE PhDs (Sanofi) and Imagine Thesis Awards. Q.R. is a recipient of an Institut Imagine MD-PhD fellowship (funded by the Fondation Bettencourt Schueller) and a Société Nationale Française de Médecine Interne (SNFMI) fellowship. We thank the LabTech Single-Cell@Imagine, supported by the Paris Region and the “Investissement d’avenir” program through 2019 ATF funding – Sésame Filières PIA (grant 3877871), for the generation of scRNA-seq librairies. We thank Imagine genomic, and bioinformatics core facilities, and the Pitié-Salpêtrière Cytometry platform CyPS for advice andtechnical assistance. We sincerely thank the patients and their families for participating in the study.

## Author contribution

Conceptualization : CC, GB, FA, MD, FRL, MMM ; Data curation: CC, TF, FC, LC; Formal analysis: CC, LD, MB, QR, CMa, GB; Investigation: CC, LD, MB, QR, ML, AR, BH, CMe, MP, MZ, BPP, VGP; Methodology: CC, GB, MMM; Funding acquisition: JCG, FA, MD, MMM; Resource: CB, EC, AB, BBM, PQDM, MLF, BN; Supervision: JCG, GB, FA, AF, MD, FRL, MMM; Visualization: CC, LD, ML, QR; Writing – original draft: CC, MMM; Writing – review & editing: CC, MLF, BN, GB, FA, AF, MD, MMM

## Declaration on interest

CC, FRL and MMM are listed as inventors on a patent application related to this article (European Patent Application no. PCT/FR2023/050433, entitled “A gene signature for diagnosing stimulator of interferon genes (STING) -associated vasculopathy with onset in infancy (SAVI)”). Other authors have no relevant interest to disclose.

## Material and methods

### Resource Availability Lead Contact

Further information and requests for resources and reagents should be directed to and will be fulfilled by the Lead Contact, Mickaël Ménager.

### Patients

Human samples preparation has been performed under the approval of the *Comité de Protection des Personnes Ile de France II* and consent for all participant has been gathered. Patients and healthy individuals participating to this study are strictly anonymized and can be identified only by members of the research team listed as author of this manuscript.

Some have been part of other studies as indicated in Figure S1A.

## Method details

### Sample collection of the SAVI dataset

Peripheral blood samples were collected on lithium heparin. Blood samples were centrifuged at 2,300 rpm (1,065g) for 10 minutes to collect supernatant (plasma), which was stored at - 20°C. Peripheral blood mononuclear cells (PBMCs) were isolated by density gradient centrifugation (2,200 rpm or 974g without break for 30 minutes) using Ficoll (Eurobio Scientific, Les Ulis, France). After centrifugation, the pellet was resuspended in Phosphate-Buffered Saline (PBS) (Thermo Fisher scientific, Illkirch, France) and cells were centrifuged at 1500 rpm (453g) for 5 minutes. Finally, the pellet was frozen in a medium containing 90% of Fetal Bovine Serum (FBS) (GIBCO, Thermo Fisher scientific, Illkirch, France) and 10% of Dimethyl Sulfoxide (DMSO) (Sigma Aldrich, St. Quentin Fallavier, France), and stored in liquid nitrogen.

### Cell phenotyping by flow cytometry

For each sample of the SAVI dataset, 500,000 cells were stained. After an initial wash and 350g centrifugation, Zombie NIR™ Fixable Viability kit was used to stain dead cells according to manufacturer instructions. Cells were washed and resuspended in 200μL of wash buffer (PBS, 5% BSA, 1mM EDTA), before staining with 90μL of antibody mix with antibodies diluted 1/50. The antibodies used in each one of the three panels are listed in supplementary file 8. Fc Block (Biolegend Europ, Netherland) and wash buffer were used to avoid unspecific bindings. For each panel, extracellular staining has been performed for 30 min at 4°C protected from light. Samples were analyzed using the Sony SP6800 spectral flow cytometers.

### IFN-β stimulation of healthy PBMC

Buffy coats from three healthy young male (19 to 22 years old) were obtained from the *Etablissement Français du Sang*. PBMCs were isolated using Ficoll (Eurobio Scientific, Les Ulis, France) density gradient and centrifugation at 800g without brakes for 25 minutes. Following centrifugation, cells were washed in wash buffer and centrifuged at 160g for 9 minutes at 4°C and the pellet was resuspended in 10mL of wash buffer. Cells were plated in 96-wells tissue culture plates, with 150,000 cells per wells in an initial volume of 125μL. One plate was made for each donor for each timepoint to plate a total of 15 million cells per condition.

Cells were challenged with 25μL of recombinant IFN-β (Human interferon beta 1a, mammalian, Catalog No 11410-2, PBL assay Science, USA) at 6,000 IU/mL for a final concentration of 1000 U/mL per well, at one of several timepoints: 1h, 2h, 4h, 8h, 12h, 24h or 36h. Stimulations were done so that all cells, even unstimulated, would be kept a total of 36h in culture. After 36h of culture at 37°C, 5% of CO2, cells were collected and counted. 10,000 cells were sent to scRNA-seq and 2 million to cell phenotyping by CyTOF. Supernatant was frozen at -20°C and kept for analysis of secreted proteins.

### Cell Phenotyping by cytometry by time-of-flight

High dimensional immune profiling of the cultured cells was obtained using the Maxpar Direct Immune Profiling System (Fluidigm, Inc France) with a 30-marker antibody panel, for CyTOF (Cytometry by Time-Of-Flight), (supplementary file 8). Briefly, 2 million cells were washed with MaxPar Cell Staining Buffer, and the cells were transferred in polypropylene tubes and incubated at room temperature for 10 minutes with 5 μL of TruStain FcX (Biolegend Europ, Netherland). Cells were then transferred into the tube containing the dry antibody cocktail, vortexed and incubated for 30 minutes at room temperature. After washing, the cells were fixed with 1mL of 1.6% paraformaldehyde solution (SigmaAldrich, France) and incubated at room temperature for 10 minutes. After washing, 1mL CellID intercalator-Ir at 125nM (pentamethylcyclopentadienyl-Ir (III)- dipyridophenazine, Fluidigm, Inc France) was added onto the cells and incubated overnight. Cells were then stored at -80°C until acquisition.

Cells were thawed at room temperature then washed 3 times in Maxpar Cell Acquisition Solution Plus (CAS+), a high-ionic-strength solution, and resuspended at a concentration of 1 million cells per mL in CAS+. 10% of EQ Four Element Calibration Beads were added to the cells immediately before acquisition. Sample acquisition and data normalization were made on the CyTOF XT mass cytometer and CyTOF software version 8.0.14050 (Fluidigm, Inc Canada) at the “Plateforme de Cytométrie de la Pitié-Salpetriere (CyPS).” An average of 500,000 events were acquired per sample. Dual count calibration, noise reduction, cell length threshold between 10 and 150 pushes, and a lower convolution threshold equal to 12 were applied during acquisition. Data cleaning was performed using Maxpar Pathsetter software v2.0.45. 4 parameters (center, offset, residual and width) were used to resolve ion fusion events (doublets) from single events using the Gaussian distribution generated by each event. After data cleaning, the program produces new FCS files consisting of only intact live single cells. Cleaned FCS files were loaded in R using the flowCore package. FCS files from all samples were concatenated into a SingleCellExperiment object. Clustering was performed using the cluster function from the CATALYST package to identify 60 initial clusters. Clusters were manually identified based on marker expression following Fluidigm Maxpar Immune Profiling manufacturer instruction. Clusters were merged into cohesive cell populations, resulting in 26 final cell types (including 3 groups of contaminants that were removed). Kinetics of cell type proportions over time was plotted using the ggplot2 package.

### scRNA-seq libraries and sequencing

scRNA-seq experiments were performed at the “Labtech SingleCell@Imagine”, in 3 batches for the SAVI dataset, and 3 batches for the IFN-β dataset. 8 libraries of the STING1 batch were generated using Chromium Single Cell 3′ Library & Gel Bead Kit v.2 (10x Genomics) according to the manufacturer’s protocol, while the remaining 9 libraries (STING2 and STING3) were generated using Chromium Single Cell 3′ Library & Gel Bead Kit v.3 (10x Genomics). Briefly, cells were counted, diluted at 1,000 cells/µL in PBS+0,04% and 20,000 cells were loaded in the 10x Chromium Controller to generate single-cell gel-beads in emulsion. After reverse transcription, gel-beads in emulsion were disrupted. Barcoded complementary DNA was isolated and amplified by PCR. Following fragmentation, end repair and A-tailing, sample indexes were added during index PCR. All libraries were sequenced on a Novaseq (Illumina). The 8 purified libraries generated from the v2 kit were sequenced with 26 cycles of read 1, 8 cycles of i7 index and 98 cycles of read 2, while the 9 libraries generated with v3 were sequenced with 28 cycles of read 1, 8 cycles of i7 index and 91 cycles of read 2. The 24 scRNA-seq libraries of the IFN-β dataset were generated using Chromium Single Cell Next GEM 3′ Library & Gel Bead Kit v.3.1 (10x Genomics) according to the manufacturer’s protocol, following the same steps as in the SAVI dataset. The purified libraries were sequenced on a Novaseq (Illumina) with 28 cycles of read 1, 8 cycles of i7 index and 91 cycles of read 2.

### Bioinformatics analysis of scRNAseq data

Sequencing reads were demultiplexed and aligned to the human reference genome (hg38), and counted using the CellRanger Pipeline v6.0. Unfiltered RNA UMI counts were loaded into Seurat v4 for quality control, data integration and downstream analyses. Apoptotic cells and empty sequencing capsules were excluded by filtering out cells with low number of features (Filters were defined based on the batch: STING1: nFeature <300; STING2 and STING3 <500; IFN-stim < 750) or a mitochondrial content higher than 20%. Data from each sample were normalized using sctransform, before batch correction using Seurat’s *FindIntegratedAnchors*, on the 3,000 most variable features. For the IFN-β dataset, for computational efficiency, integration anchors were determined with a canonical correlation analysis, using all unstimulated samples as reference. For the SAVI dataset, to avoid over-aligning samples, a reciprocal principal component analysis (PCA) was used to find integration anchors. On the resulting integrated datasets, we computed the PCA on the 3000 most variable genes, before computing a UMAP (on 20 PCs for SAVI and 30 PCs for IFN-stimulation). Community detection was performed using the graph-based modularity-optimization Louvain algorithm from Seurat’s *FindClusters* function with a 1.2 resolution. Cell types labels were assigned to resulting clusters based on a manually curated list of marker genes as well as previously defined signatures of the PBMC subtypes.^58^ Despite filtering for high quality cells, 11/34 clusters in the SAVI dataset and 7/36 in the IFN-β dataset stood out as poor quality clusters and were removed from further analysis. They represented low UMI cells, dying cells, cycling cells, doublets, or contaminants (progenitor cells, basophils, platelets or megakaryocytes). A total of 112,060 and 115,503 cells were kept in the SAVI and IFN-β datasets respectively. Transcriptomic signatures scores were calculated using Seurat’s *AddModuleScore* function. The signatures and their origin are described in supplementary file 8. Differential expression testing was conducted using the *FindMarkers* function in Seurat, with default Wilcoxon testing. *P*-values were controlled using Bonferroni correction. Genes with an absolute log(fold-change) ≥0.25 and an adjusted *p*-value ≤0.05 were selected as differentially expressed. Pathway analysis was performed using both the Ingenuity pathway analysis v57662101 software (IPA (QIAGEN Inc.) and EnrichR ^59, 60^. Heatmaps were extracted from the comparison module in IPA. Pathways with an absolute z-score lower than 2 or a Bonferroni-Hochberg corrected p-values higher than 0.05 were filtered out. The TRRUST transcription factors 201921 used for the transcription factors enrichment analysis was performed using Enrich R.

The cell cycle phase was defined using the *CellCycleScoring* function of Seurat V4, with the default gene lists and adjusting the parameters to better reflect expected proportions of CTRL cells in each phase of the cell cycle.

Cell-to-cell interactions were predicted using the ICELLNET framework^26^. We filtered out filtered the gene expression matrix to only keep the genes that were expressed in at least 5% of the cells in at least one of the clusters. We combined together cluster 5 and cluster 17 into a single classical monocyte group and considered this group as the “central cells”. Each cluster of CD4^+^ or CD8^+^ T cell was considered as the “partner cells”. In the SAVI dataset, the predictions were performed separately for the CTRLs, the SAVI, and the SAVI_treated groups. In the IFN-β dataset, the predictions were performed separately for each timepoint.

### Cytokine measurements

The plasma samples and conditioned media from in vitro stimulated PBMC were analyzed by proximity extension assays (PEA), a targeted, antibody-based, proteomics method. We used the PEA Olink Target96 Inflammation panel which targets 92 biomarkers (supplementary file 8). Briefly, 2 DNA-labeled antibodies bind to a targeted protein, followed by hybridization of the oligonucleotides when the antibodies come in close proximity to each other. Adding DNA polymerase to the sample generates a unique PCR target sequence, which is then detected and quantified using a quantitative PCR (qPCR) readout (BiomarkHD™, Data collection and Real Time PCR analysis software 4.7.1, Fluidigm).

The Olink PEA Quality control process consists of technical controls to monitor the performance of all 3 steps of the assays (immunoreaction, extension, and amplification/detection) as well as the individual samples. Internal controls are spiked into each sample to provide a non-human assay control, a positive extension control consisting of an antibody coupled to a unique DNA-pair and, finally, a positive detection control based on a double stranded DNA amplicon. In addition, each experimental run includes a control strip with control samples used to estimate the accuracy of protein detection and quantification (intra- and inter-coefficient of variation). A negative control (buffer) run in triplicate is used to set background levels and calculate limit of detection (LOD), a plate control (plasma pool) is run in triplicate for plate normalization, and a sample control (reference plasma) is included in duplicate to estimate CV between runs.

Data are presented as NPX (Normalized Protein eXpression) values. NPX is Olink’s relative protein quantification unit on log2 scale. Data generation of NPX (obtained using Olink NPX Manager 3.3.2.434) consists of normalization to the extension control (known standard), log2- transformation, and level adjustment using the plate control (plasma sample).

Serial dilutions of samples are performed to determine a potential Hook effect (observed when there is an antigen excess relative to the reagent antibodies, resulting in falsely lower values). Conditioned media are tested at 1:1, 1:4 and 1:10 dilutions for Target96 Inflammation. Plasma samples are not diluted.

For plasma cytokine measurement, differential protein expression was performed using Multiple comparisons parametric test (TIBCO® Spotfire® v.10.3.2.7) with Benjamini-Hochberg (BH) False Discovery Rate adjustment. For the IFN-β dataset, pairwise analysis by donor was performed by timepoint. For patient samples, SAVI samples were compared to healthy donors. An adjusted *p*-value<0.05 was considered statistically significant.

### Exome analysis

The exome sequencing used in this study has been described by *Jeremiah et al* ^1^. Exome sequencing was performed by the Centre National de Génotypage, Institut de Génomique, CEA. After quality control by the DNA Bank Laboratory, genomic DNA (3µg) was captured using in-solution enrichment methodology (Human All Exon v5 – 50 Mb, Agilent Technologies, CA, USA). Library preparation and exome enrichment protocol (∼20,000 targeted genes) was performed on an automated platform, using NGSx (Perkin Elmer Inc, MA, USA) and Bravo (Agilent Technologies, CA, USA) robots respectively, according to manufacturer’s instructions (SureSelect, Agilent Technologies CA, USA). After normalization and quality control, exome enriched libraries were sequenced on a HiSEQ 2000 (Illumina Inc., CA, USA) as paired-end 100 base reads. Sequencing was performed in order to provide a mean cover of at least 60 to 70X for each sample. Image analysis and base calling was performed using Illumina Real Time Analysis (RTA) Pipeline. Sequence quality parameters were assessed daily throughout the 12 days sequencing run. Sequences were aligned to the reference human genome hg19 using the Burrows-Wheeler Aligner. Downstream processing was carried out with the Genome Analysis Toolkit (GATK), SAMtools, and Picard, following documented best practices (http://www.broadinstitute.org/gatk/guide/topic?name=best-practices).

We searched for SNPs in the 85 genes of the UPR signature (supplementary file 8), to extract the SNPs found in less than 5% of the general population. No other filter was used.

We generate a HOPE report (https://www3.cmbi.umcn.nl/hope/)^61^ for the p.P649T mutation in PERK (encoded by *EIF2AK3*), by using PERK uniprot identifier Q9NZJ5.

### Generation of the ADUS100 dataset

Human PBMC samples were obtained from Promocell (C-12907 promocell, lot 434Z036). Briefly, after thawing and a quality control on cell viability (>90%), a minimal of of 2*10^6^ cells cells were incubated either with or without ADUS100 (3mM final concentration), a STING agonist, during 4h in RPMI1640 medium complemented with 10% heat inactivated fetal bovine serum. In some conditions, cells were pre-incubated 15min with a JAK1-2 inhibitor (baricitinib, 10mM final concentration) prior and during ADUS100 exposure. After 4h, cells were washed with PBS without Ca/Mg containing 0.04% BSA and a minimum of 6,105 harvested for single cell analysis (10X Genomics). Libraries were generated using Chromium Single Cell 3′ Library & Gel Bead Kit v.2 (10x Genomics) according to the manufacturer’s protocol. Libraries were sequenced on a Novaseq (Illumina), with 26 cycles of read 1, 8 cycles of i7 index and 98 cycles of read 2.

### Primary and SV40-fibroblasts

Patient’s primary fibroblasts were obtained by cultivating skin biopsies in RPMI 1640 supplemented with 20% FBS, 1% penicillin/streptomycin, 1X gentamicin, and 1X amphotericin B. Primary fibroblasts were transformed by lipofection of pBSSVD2005 (Addgene #21826) coding the large-T antigen of Simian virus-40. After 3 to 4 cycles of cultures, fibroblasts change of their shape and growing rate and were considered immortalized. This was later confirmed by evaluating expression of SV40 by confocal microscopy. SV40- fibroblasts were then maintained in DMEM supplemented with 10% FBS and 1% penicillin/streptomycin.

### Sanger sequencing

Sanger sequencing was performed on SV40-fibroblast genomic DNA to confirm the next-generation sequencing results for *EIF2AK3*. The primers used for PCR are as follow: Forward TCAGATATGAACAGCCTTCAGTGT; Reverse AACCAAAATTTCACAAGTGGCT.

Purified PCR products were directly sequenced using BigDye Terminators (version 1.1) and a 3500xL Genetic Analyzer (Applied Biosystems).

## Quantification and statistical analysis

Statistical tests for cellular composition analysis in both the CyTOF and scRNA-seq datasets were performed in R v3.6.1. Kruskal-Wallis test followed by post-hoc multiple comparison Dunn’s test was applied to assess differences in cell population proportions (*: *p* ≤0.05; **: *p* ≤0.01; ***: *p* ≤0.001).

## Supplementary information

• Supplementary 1: SAVI dataset – DEGs in the main populations, related to Figure 2

• Supplementary 2: IFN dataset – DEGs of 36h vs 0h, in the main populations, related to Figure 2

• Supplementary 3: IFN dataset – DEGs of each timepoint vs 0h, in all PBMCs, related to Figure S2

• Supplementary 4: SAVI dataset – DEGs of SAVI vs CTRLs, in each T cell cluster, related to Figure 3

• Supplementary 5: SAVI dataset – DEGs of cluster 17 vs cluster 5, in the SAVI group, related to Figure 4

• Supplementary 6: SAVI dataset – DEGs in each monocyte cluster, related to Figure S4

• Supplementary 7: STING activation signature, related to Figure 6

• Supplementary 8: Methods

---

**Figure S1:**
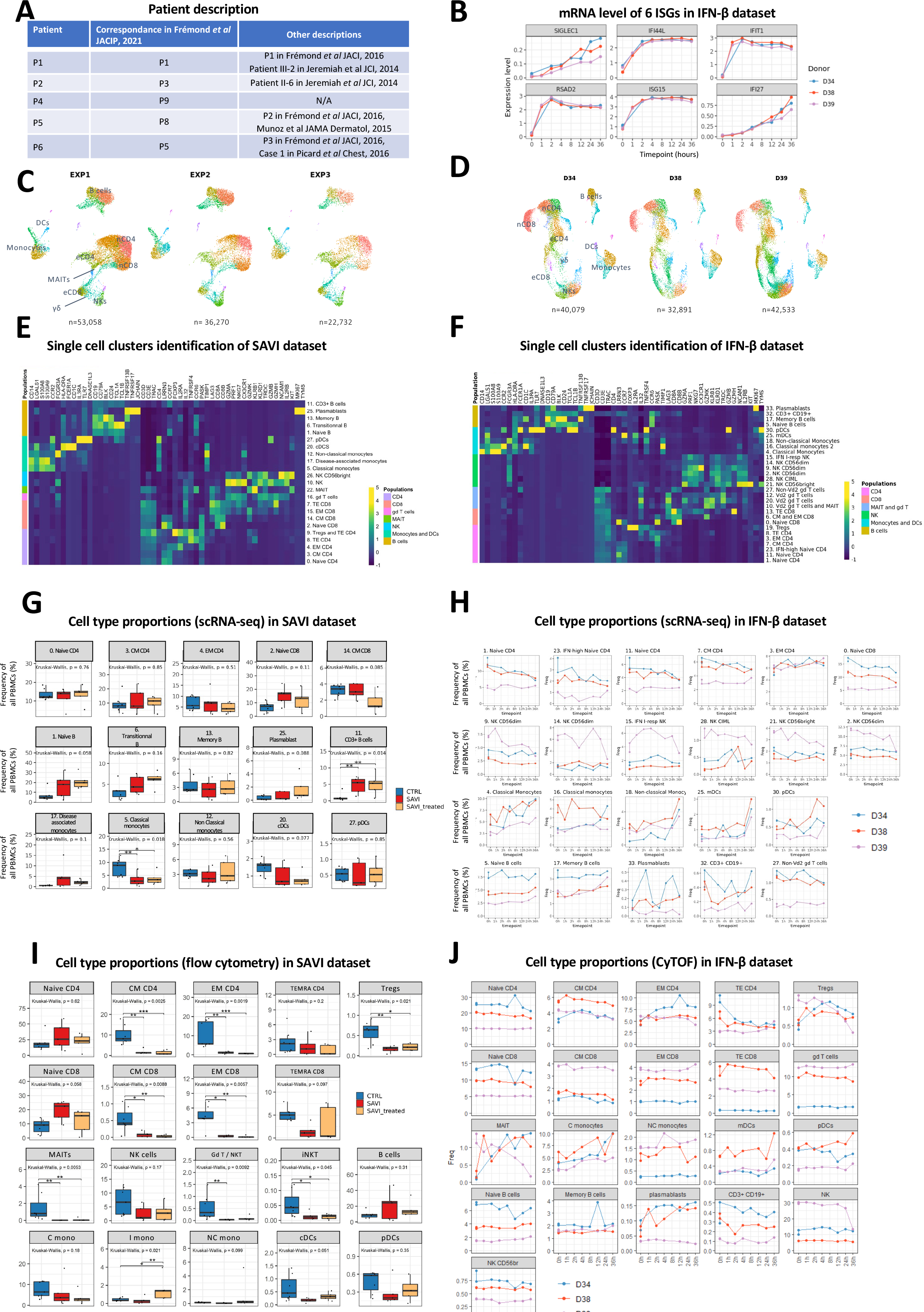
scRNA-seq and phenotyping of the SAVI dataset and the IFN-β dataset A. The identification number of the SAVI patients in Frémond *et al*, 2021 B. IFN-β dataset: Validation of the IFN-β stimulation using the mRNA levels measured by scRNA-seq of 6 well known IFN-stimulated genes C. UMAP of the SAVI dataset separated by batch. The number of cells in each group is indicated below the UMAP. nCD4: naïve CD4; eCD4: effector CD4; nCD8: naïve CD8; eCD8: effector CD8; γδ: γδ T cells D. UMAP of the IFN-β dataset separated by donor. The number of cells in each group is indicated below the UMAP. nCD4: naïve CD4; eCD4: effector CD4; nCD8: naïve CD8; eCD8: effector CD8; γδ: γδ T cells E. Heatmap of the average expression of cell type markers in each cluster of the SAVI dataset. These markers were used to manually assign a cell type to each cluster F. Heatmap of the average expression of cell type markers in each cluster of the IFN-β dataset. These markers were used to manually assign a cell type to each cluster. G. Boxplot of the proportion of PBMCs found each cluster of the SAVI dataset, by scRNA-seq. *p*-values are calculated by Kruskal-Wallis test for multiple comparisons, followed by a post hoc Dunn’s test. *(*p* < 0.05), **(*p* < 0.01), ***(*p* < 0.001) H. Evolution of the proportion of PBMCs found in each cluster of the IFN-β dataset over the time course of IFN-β stimulation, by scRNA-seq I. Boxplot of the proportion of PBMCs found each cell population of the SAVI dataset, by flow cytometry. *p*-values are calculated by Kruskal-Wallis test for multiple comparisons, followed by a post hoc Dunn’s test. *(*p* < 0.05), **(*p* < 0.01), ***(*p* < 0.001). C mono: classical monocytes; I mono : intermediary monocytes; NC mono : non-classical monocytes. J. Evolution of the proportion of PBMCs found in each cluster of the IFN-β dataset over the time course of IFN-β stimulation, by Cytometry by Time of Flight

**Figure S2:**
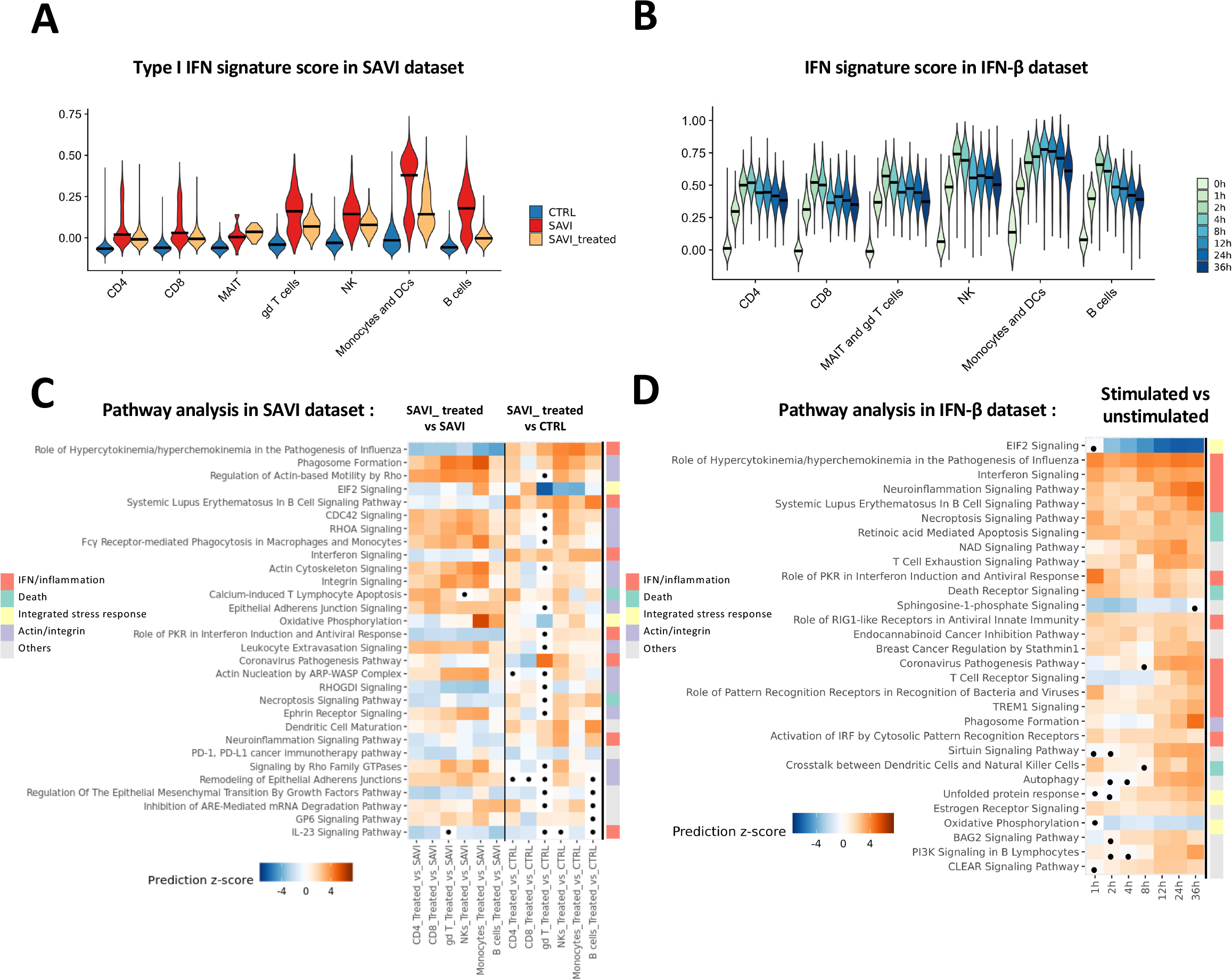
Pathway analysis in the SAVI dataset and the IFN-β dataset. A. Violin plot of a type I IFN response signature of 272 genes, in the SAVI dataset. Dark lines indicate medians B. Violin plot of a type I IFN response signature of 272 genes, in the IFN-β dataset. Dark lines indicate medians C. Heatmap of the pathway enrichment analysis, performed in IPA, between SAVI_treated and SAVI and between SAVI_treated and CTRL in each cell compartment. Dots indicate non-significant pathways (Bonferroni-Hochberg corrected p-values > 0.05). Side color bar indicate groups of pathways based on broader functions. Color scales indicate direction of prediction with orange for predicted activation and blue for predicted inhibition D. Heatmap of the pathway enrichment analysis, performed in IPA, between PBMCs stimulated with IFN-β for each timepoint and unstimulated PBMCs. Dots indicate non-significant pathways (Bonferroni-Hochberg corrected p-values > 0.05). Side color bar indicate groups of pathways based on broader functions. Color scales indicate direction of prediction with orange for predicted activation and blue for predicted inhibition

**Figure S3:**
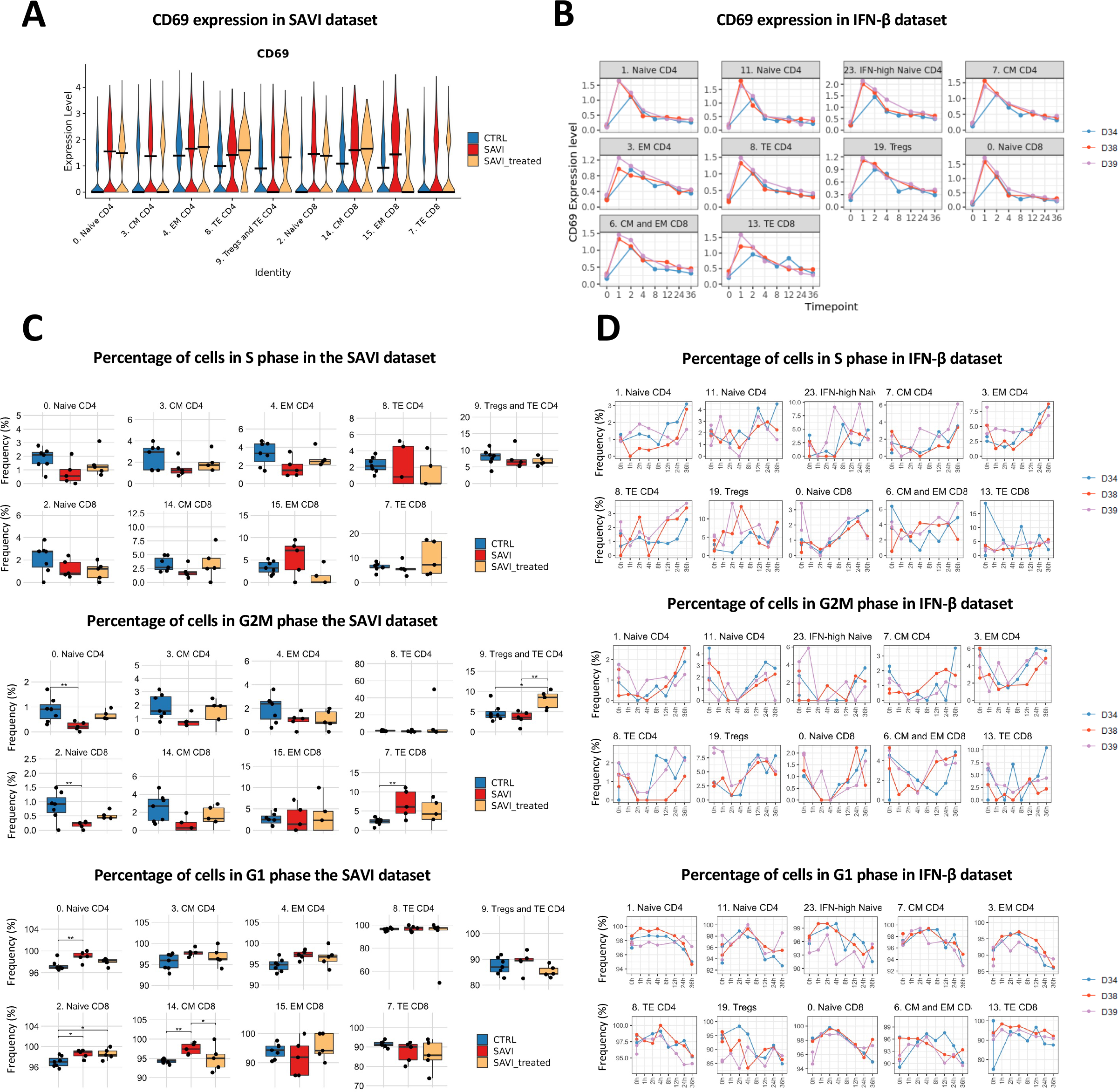
*CD69* expression and cell cycle phase at the transcriptomic level in CD4^+^ and CD8^+^ T cells the SAVI dataset and the IFN-β dataset A. Violin plot of *CD69* expression, in CD4^+^ and CD8^+^ T cells of the SAVI dataset. Dark lines indicate medians B. Evolution of *CD69* expression in CD4^+^ and CD8^+^ T cells of the IFN-β dataset over the time course of IFN-β stimulation. Each dot is the average score of the signature for a sample C. Boxplot of the proportion of cells in the S phase (top), the G2M phase (middle), and the G1 phase (bottom) of the cell cycle, in CD4^+^ and CD8^+^ T cells of the SAVI dataset D. Evolution of the proportion of cells in the S phase (top) the G2M phase (middle), and the G1 phase (bottom) of the cell cycle in CD4^+^ and CD8^+^ T cells of the IFN-β dataset over the time course of IFN-β stimulation

**Figure S4:**
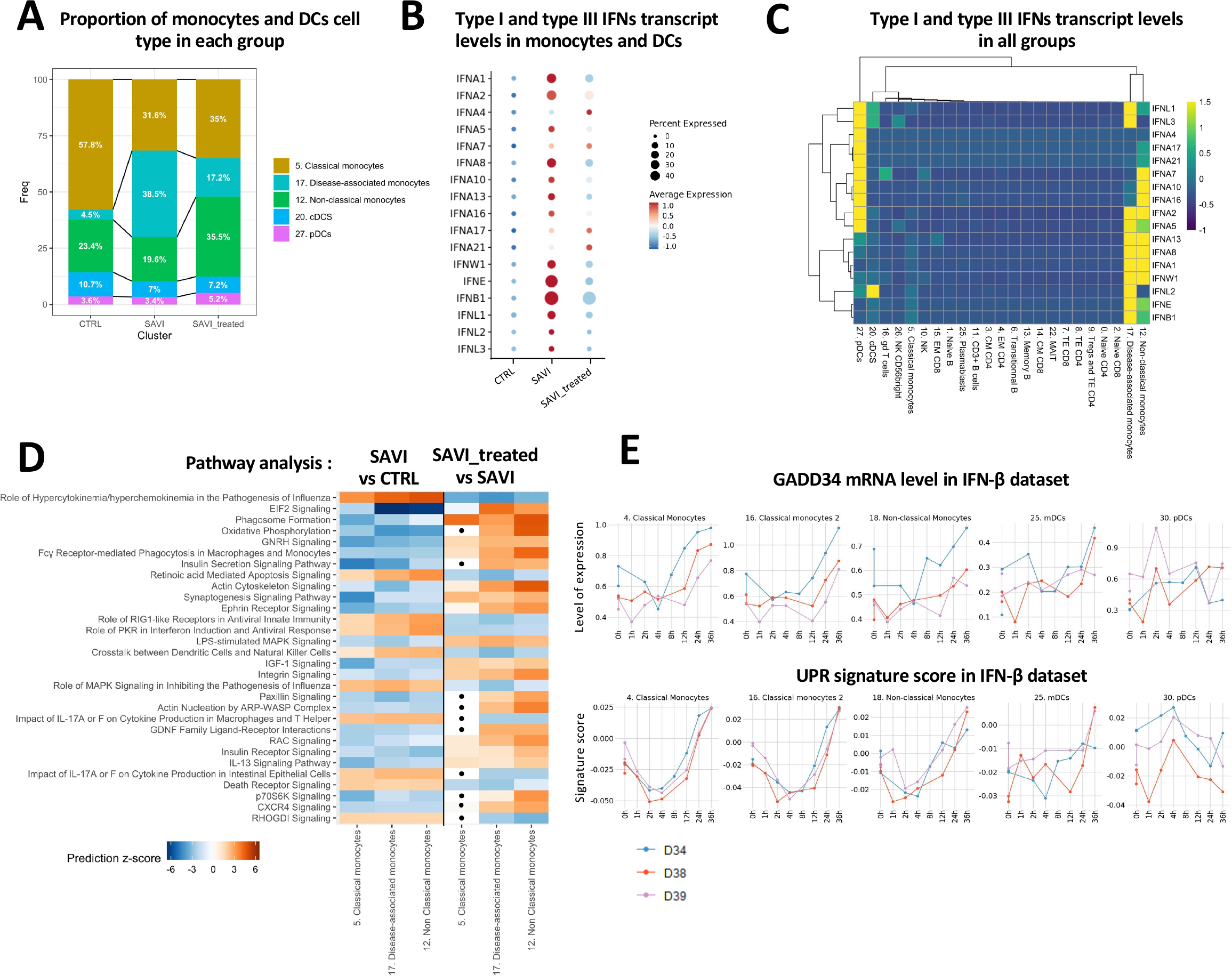
Further characterization of a disease-associated cluster of monocytes at the transcriptomic level A. Proportion of cells from each monocyte and dendritic cell clusters in each group in the SAVI dataset B. Dot plot of the scaled average expression of each type I and type III IFN in all monocytes and DCs clusters in each group of the SAVI dataset. Dot size indicates the percentage of cells expressing the gene and color scale indicates the average expression of the gene in each group. C. Heatmap of the scaled expression levels of all type I and type III IFN, in each cluster, in all cells of the SAVI dataset. Hierarchical clustering based on Euclidian distances. D. Heatmap of the pathway enrichment analysis, performed in IPA, between SAVI and CTRL, and between SAVI_treated and SAVI in the clusters of the monocyte compartment. Dots indicate non-significant pathways (BH > 0.05) E. Evolution of the expression of *PPP1R15A* which codes for GADD34 (left) and of an unfolded protein response signature of 85 genes (right) in each monocyte or dendritic cell cluster of the IFN-β dataset over the time course of IFN-β stimulation. Each dot is the average score for a sample

**Figure S5:**
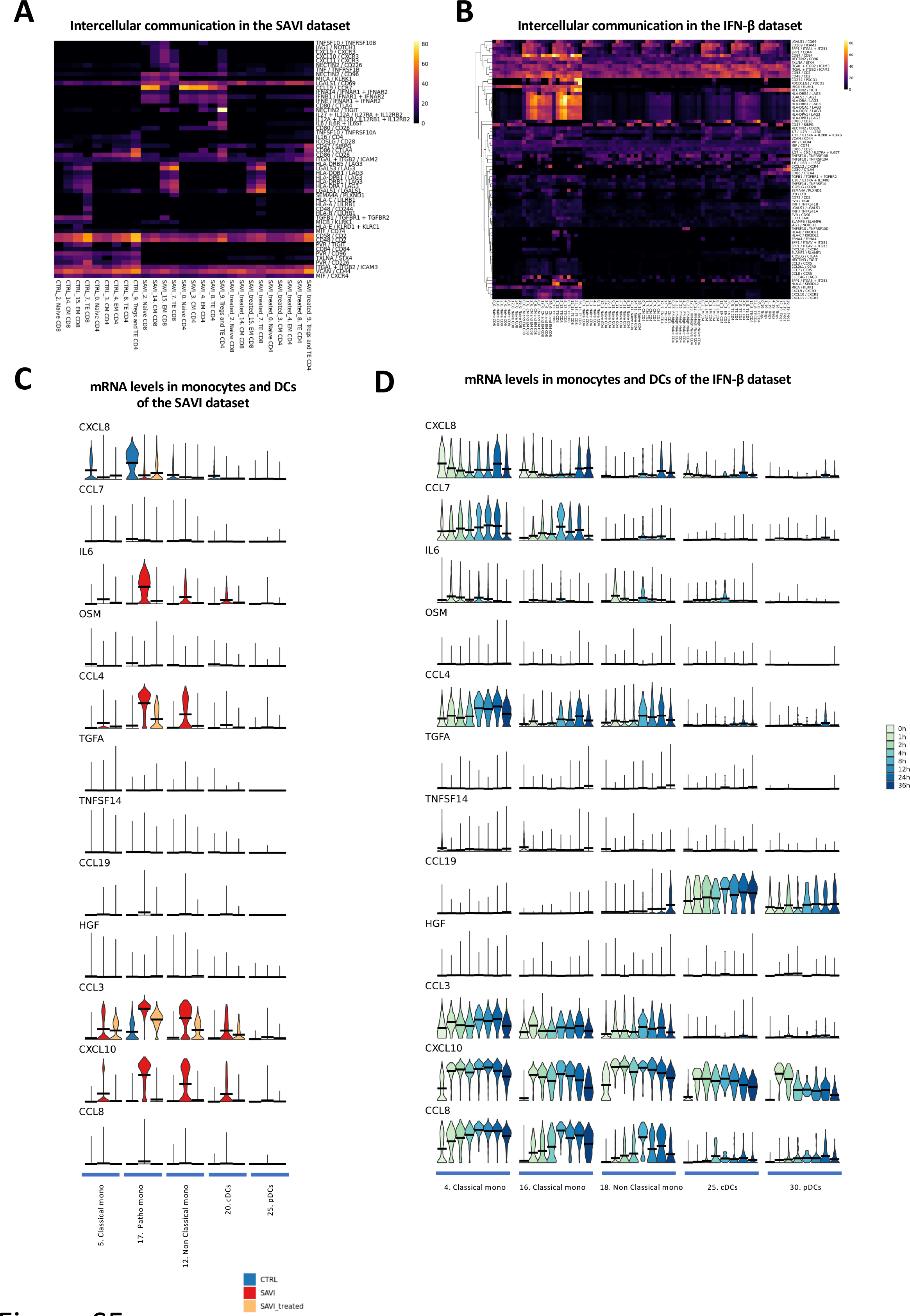
Ligand/receptors prediction and mRNA levels of secreted proteins A. Heatmap of the score of each ligand/receptor pair between each T cell cluster and the monocytes, either in the SAVI, the treated_SAVI or the CTRL group B. Heatmap of the score of each ligand/receptor pair between each T cell cluster and the monocytes, for each timepoint of IFN-β stimulation. Hierarchical clustering based on Pearson correlation C. Violin plot of the mRNA levels of the 12 proteins found upregulated in the blood of SAVI patients in Figure 5F, in each monocyte and DC clusters of the SAVI dataset D. Violin plot of the mRNA levels of the 12 proteins found upregulated in the blood of SAVI patients in Figure 5F, in each monocyte and DC clusters of the IFN-β dataset

**Figure S6:**
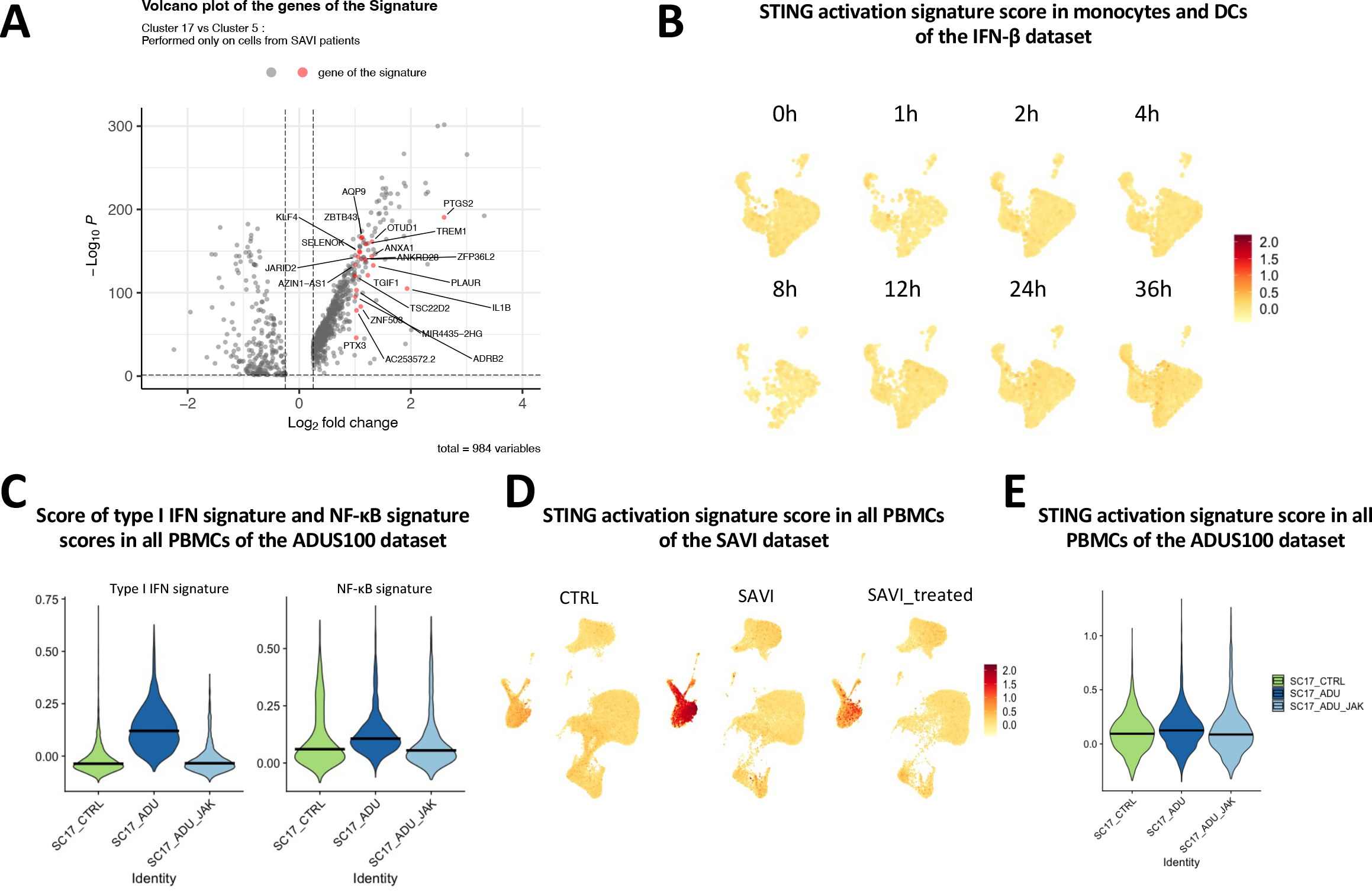
Design of a STING-activation signature, independent of type I IFN response A. Volcano plot of the DEGs between cluster 17 and cluster 5, in SAVI, with the 21 genes of the STING activation signature highlighted in red. B. Feature plot of the signature score of the 21 genes of the STING activation signature in the monocytes and DCs of each timepoint of the IFN-β dataset C. Violin plots of the score of a type I IFN response signature of 272 genes and of an NF-κB activation signature of 200 genes in all PBMCs of CTRL, ADUS100 and ADUS100+JAK-inhibitor in the ADUS100 dataset. Dark lines indicate medians D. Feature plot of the signature score of the 21 genes of the STING activation signature in all PBMCs of CTRL, SAVI and SAVI_treated in the SAVI dataset E. Violin plot of the signature score of the 21 genes of the STING activation signature in all PBMCs of CTRL, ADUS100 and ADUS100+JAK-inhibitor in the ADUS100 dataset. Dark lines indicate medians

**Figure S7:**
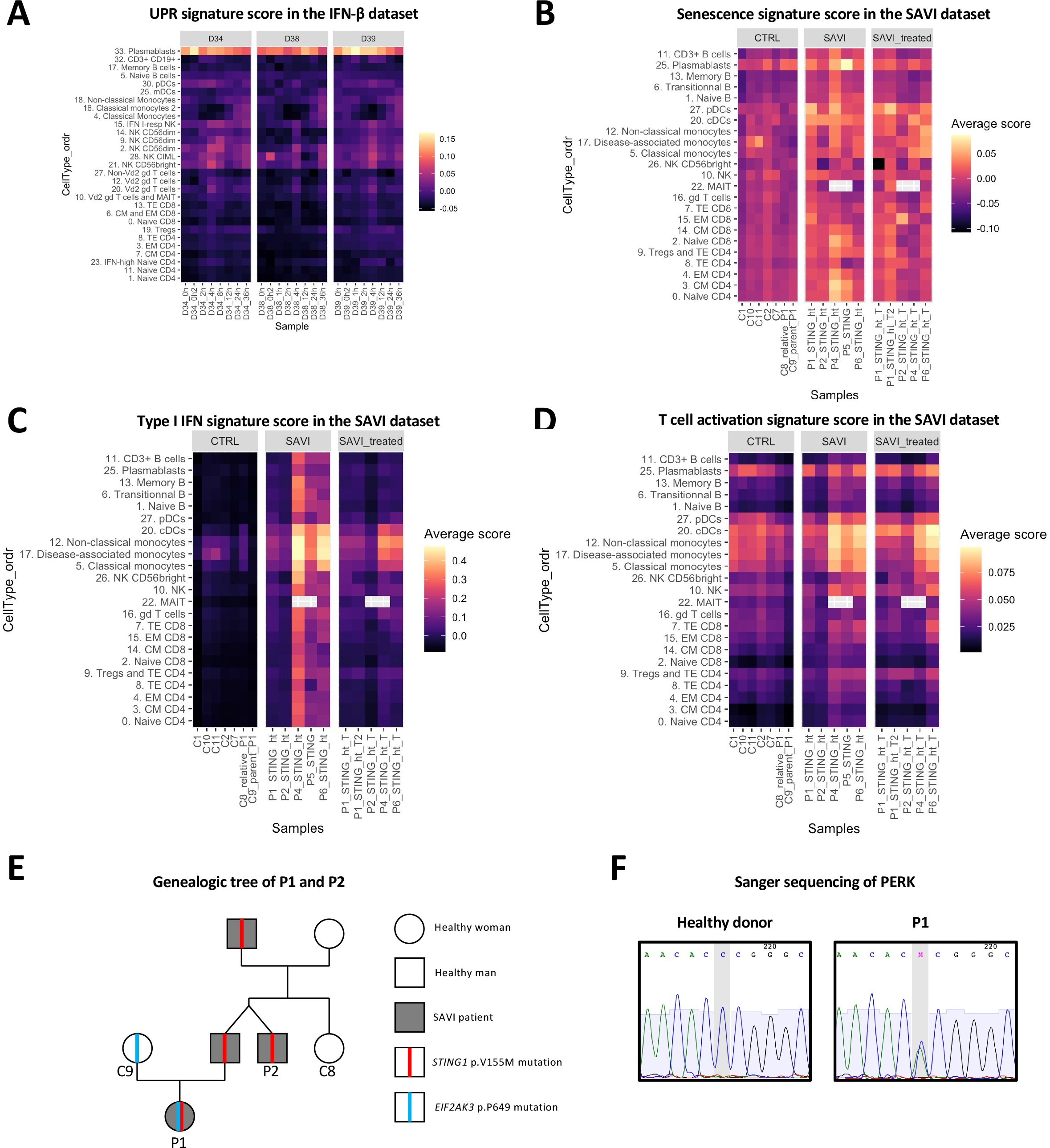
Transcriptomic profiles of signatures relevant to P1 A. Heatmap of a UPR signature of 85 genes in each sample and each cluster of the IFN-β dataset B. Heatmap of a senescence signature of 50 genes in each sample and each cluster of the SAVI dataset. Grey squares are used when no cells from a sample in found in a cluster. C. Heatmap of a type I IFN response signature of 272 genes in each sample and each cluster of the SAVI dataset. Grey squares are used when no cells from a sample in found in a cluster. D. Heatmap of a T cell activation signature of 4917 genes in each sample and each cluster of the SAVI dataset. Grey squares are used when no cells from a sample in found in a cluster. E. Genealogic tree of P1’s family. Squares represent male individuals and rounds represent female individuals. F. Sanger sequencing of genomic DNA from SV40-fibroblasts of a healthy control and P1. The « M » stands for a heterozygous C>A variation.

## Notes

### Funding Statement

The study was supported by the institut National de la Santé et de la Recherche Médicale (INSERM), by the AtipAvenir, by government grants managed by the Agence Nationale de la Recherche as part of the Investment for the future program (Institut Hospital universitaire Imagine, grant ANR 10 IAHU01, Recheche Hospitalo Universitaire, grant ANR 18 RHU S622 0010) by a CIFRE PhD program with Sanofi iAward Europe, and the Emergence ville de Paris program. C.C and L.D. are both the recipients of CIFRE PhDs (Sanofi) and Imagine Thesis Awards. Q.R. is a recipient of an Institut Imagine MD PhD fellowship (funded by the Fondation Bettencourt Schueller) and a Société Nationale Française de Médecine Interne (SNFMI) fellowship. The LabTech SingleCell@Imagine is supported by the Paris Region and the Investissement d avenir program through 2019 ATF funding Sésame Filières PIA (grant 630 3877871)

### Author Declarations

Comité de Protection des Personnes Ile de France II gave ethical approval for this work

## References

1. Jeremiah, N. et al. Inherited STING-activating mutation underlies a familial inflammatory syndrome with lupus-like manifestations. J Clin Invest 124, 5516–5520 (2014).

2. Liu, Y. et al. Activated STING in a Vascular and Pulmonary Syndrome. New England Journal of Medicine 371, 507–518 (2014).

3. Frémond, M.-L. et al. Overview of STING-Associated Vasculopathy with Onset in Infancy (SAVI) Among 21 Patients. The Journal of Allergy and Clinical Immunology: In Practice 9, 803–818.e11 (2021).

4. David, C. & Frémond, M.-L. Lung Inflammation in STING-Associated Vasculopathy with Onset in Infancy (SAVI). Cells 11, 318 (2022).

5. Dobbs, N. et al. STING Activation by Translocation from the ER Is Associated with Infection and Autoinflammatory Disease. Cell Host & Microbe 18, 157–168 (2015).

6. Ishikawa, H., Ma, Z. & Barber, G. N. STING regulates intracellular DNA-mediated, type I interferon-dependent innate immunity. Nature 461, 788–792 (2009).

7. Abe, T. & Barber, G. N. Cytosolic-DNA-mediated, STING-dependent proinflammatory gene induction necessitates canonical NF-κB activation through TBK1. J Virol 88, 5328–5341 (2014).

8. Yum, S., Li, M., Fang, Y. & Chen, Z. J. TBK1 recruitment to STING activates both IRF3 and NF-κB that mediate immune defense against tumors and viral infections. Proc Natl Acad Sci U S A 118, e2100225118 (2021).

9. Tanaka, Y. & Chen, Z. J. STING Specifies IRF3 phosphorylation by TBK1 in the Cytosolic DNA Signaling Pathway. Sci Signal 5, ra20 (2012).

10. Dou, Z. et al. Cytoplasmic chromatin triggers inflammation in senescence and cancer. Nature 550, 402–406 (2017).

11. Wu, J. et al. STING-mediated disruption of calcium homeostasis chronically activates ER stress and primes T cell death. J Exp Med 216, 867–883 (2019).

12. Gulen, M. F. et al. Signalling strength determines proapoptotic functions of STING. Nat Commun 8, 427 (2017).

13. Staels, F. et al. Adult-Onset ANCA-Associated Vasculitis in SAVI: Extension of the Phenotypic Spectrum, Case Report and Review of the Literature. Front Immunol 11, 575219 (2020).

14. Cerboni, S. et al. Intrinsic antiproliferative activity of the innate sensor STING in T lymphocytes. Journal of Experimental Medicine 214, 1769–1785 (2017).

15. Picard, C. et al. Severe Pulmonary Fibrosis as the First Manifestation of Interferonopathy (TMEM173 Mutation). Chest 150, e65–e71 (2016).

16. Tang, X. et al. STING-Associated Vasculopathy with Onset in Infancy in Three Children with New Clinical Aspect and Unsatisfactory Therapeutic Responses to Tofacitinib. J Clin Immunol 40, 114–122 (2020).

17. Lin, B. et al. A novel STING1 variant causes a recessive form of STING-associated vasculopathy with onset in infancy (SAVI). Journal of Allergy and Clinical Immunology 146, 1204–1208.e6 (2020).

18. Frémond, M.-L. et al. Efficacy of the Janus kinase 1/2 inhibitor ruxolitinib in the treatment of vasculopathy associated with TMEM173-activating mutations in 3 children. Journal of Allergy and Clinical Immunology 138, 1752–1755 (2016).

19. Gao, K. M., Motwani, M., Tedder, T., Marshak-Rothstein, A. & Fitzgerald, K. A. Radioresistant cells initiate lymphocyte-dependent lung inflammation and IFNγ-dependent mortality in STING gain-of-function mice. Proc Natl Acad Sci U S A 119, e2202327119 (2022).

20. Zhang, D. et al. A non-canonical cGAS–STING–PERK pathway facilitates the translational program critical for senescence and organ fibrosis. Nat Cell Biol 24, 766–782 (2022).

21. Munoz, J. et al. Stimulator of Interferon Genes–Associated Vasculopathy With Onset in Infancy: A Mimic of Childhood Granulomatosis With Polyangiitis. JAMA Dermatol 151, 872 (2015).

22. Picard, C. et al. Severe Pulmonary Fibrosis as the First Manifestation of Interferonopathy (TMEM173 Mutation). Chest 150, e65–71 (2016).

23. Balka, K. R. et al. TBK1 and IKKε Act Redundantly to Mediate STING-Induced NF- κB Responses in Myeloid Cells. Cell Reports 31, 107492 (2020).

24. Pakos-Zebrucka, K., et al. The integrated stress response. EMBO Rep 17, 1374–1395 (2016).

25. Reich, S. et al. A multi-omics analysis reveals the unfolded protein response regulon and stress-induced resistance to folate-based antimetabolites. Nat Commun 11, 2936 (2020).

26. Noël, F. et al. Dissection of intercellular communication using the transcriptome- based framework ICELLNET. Nat Commun 12, 1089 (2021).

27. Yan, Y. et al. CCL19 and CCR7 Expression, Signaling Pathways, and Adjuvant Functions in Viral Infection and Prevention. Front Cell Dev Biol 7, 212 (2019).

28. Shrager, S. H. & Kiel, C. SnapShot: APC/T Cell Immune Checkpoints. Cell 183, 1142–1142.e1 (2020).

29. Grebinoski, S. et al. Autoreactive CD8+ T cells are restrained by an exhaustion-like program that is maintained by LAG3. Nat Immunol 23, 868–877 (2022).

30. Cedeno-Laurent, F. & Dimitroff, C. J. Galectin-1 research in T cell immunity: Past, present and future. Clin Immunol 142, 107–116 (2012).

31. Fulda, S. Tumor-necrosis-factor-related apoptosis-inducing ligand (TRAIL). Adv Exp Med Biol 818, 167–180 (2014).

32. 32. Guerra, N. & Lanier, L. L. Editorial: Emerging Concepts on the NKG2D Receptor- Ligand Axis in Health and Diseases. Frontiers in Immunology 11, (2020).

33. Thim-Uam, A. et al. STING Mediates Lupus via the Activation of Conventional Dendritic Cell Maturation and Plasmacytoid Dendritic Cell Differentiation. iScience 23, 101530 (2020).

34. Gable, D. L. et al. ZCCHC8, the nuclear exosome targeting component, is mutated in familial pulmonary fibrosis and is required for telomerase RNA maturation. Genes Dev 33, 1381–1396 (2019).

35. Kieper, W. C. & Jameson, S. C. Homeostatic expansion and phenotypic conversion of naïve T cells in response to self peptide/MHC ligands. Proceedings of the National Academy of Sciences 96, 13306–13311 (1999).

36. Ramanathan, S. et al. Cytokine synergy in antigen-independent activation and priming of naive CD8+ T lymphocytes. Crit Rev Immunol 29, 219–239 (2009).

37. Frisch, S. M. & MacFawn, I. P. Type I interferons and related pathways in cell senescence. Aging Cell 19, e13234 (2020).

38. Yu, Q. et al. DNA-damage-induced type I interferon promotes senescence and inhibits stem cell function. Cell Rep 11, 785–797 (2015).

39. Yang, H., Wang, H., Ren, J., Chen, Q. & Chen, Z. J. cGAS is essential for cellular senescence. Proceedings of the National Academy of Sciences 114, E4612–E4620 (2017).

40. Betts, M. J. & Russell, R. B. Amino acid properties and consequences of substitutions. in Bioinformatics for Geneticists.

41. Ma, Y., Shimizu, Y., Mann, M. J., Jin, Y. & Hendershot, L. M. Plasma cell differentiation initiates a limited ER stress response by specifically suppressing the PERK- dependent branch of the unfolded protein response. Cell Stress Chaperones 15, 281–293 (2010).

42. Zhu, H. et al. Ufbp1 promotes plasma cell development and ER expansion by modulating distinct branches of UPR. Nat Commun 10, 1084 (2019).

43. Zhou, Z. et al. Phenotypic and functional alterations of pDCs in lupus-prone mice. Sci Rep 6, 20373 (2016).

44. Liao, X. et al. Cutting Edge: Plasmacytoid Dendritic Cells in Late-Stage Lupus Mice Defective in Producing IFN-α. J Immunol 195, 4578–4582 (2015).

45. Rodero, M. P. et al. Detection of interferon alpha protein reveals differential levels and cellular sources in disease. J Exp Med 214, 1547–1555 (2017).

46. Affandi, A. J. et al. CD169 Defines Activated CD14+ Monocytes With Enhanced CD8+ T Cell Activation Capacity. Frontiers in Immunology 12, (2021).

47. Théry, C. & Amigorena, S. The cell biology of antigen presentation in dendritic cells. Curr Opin Immunol 13, 45–51 (2001).

48. Wang, S. & El-Deiry, W. S. TRAIL and apoptosis induction by TNF-family death receptors. Oncogene 22, 8628–8633 (2003).

49. Roan, F., Obata-Ninomiya, K. & Ziegler, S. F. Epithelial cell-derived cytokines: more than just signaling the alarm. J Clin Invest 129, 1441–1451 (2019).

50. Misharin, A. V. et al. Monocyte-derived alveolar macrophages drive lung fibrosis and persist in the lung over the life span. J Exp Med 214, 2387–2404 (2017).

51. McCubbrey, A. L. et al. Deletion of c-FLIP from CD11bhi Macrophages Prevents Development of Bleomycin-induced Lung Fibrosis. Am J Respir Cell Mol Biol 58, 66–78 (2018).

52. Fraser, E. et al. Multi-Modal Characterization of Monocytes in Idiopathic Pulmonary Fibrosis Reveals a Primed Type I Interferon Immune Phenotype. Frontiers in Immunology 12, (2021).

53. Kessler, N. Monocyte-derived macrophages aggravate pulmonary vasculitis via cGAS/STING/IFN-mediated nucleic acid sensing. 51 (2022).

54. Barabutis, N. Unfolded Protein Response in Lung Health and Disease. Front Med (Lausanne) 7, 344 (2020).

55. Derisbourg, M. J., Hartman, M. D. & Denzel, M. S. Modulating the integrated stress response to slow aging and ameliorate age-related pathology. Nat Aging 1, 760–768 (2021).

56. Kim, D. et al. Targeted therapy guided by single-cell transcriptomic analysis in drug- induced hypersensitivity syndrome: a case report. Nat Med 26, 236–243 (2020).

57. Gaidt, M. M. et al. The DNA Inflammasome in Human Myeloid Cells Is Initiated by a STING-Cell Death Program Upstream of NLRP3. Cell 171, 1110–1124.e18 (2017).

58. Monaco, G. et al. RNA-Seq Signatures Normalized by mRNA Abundance Allow Absolute Deconvolution of Human Immune Cell Types. Cell Rep 26, 1627–1640.e7 (2019).

59. Chen, E. Y. et al. Enrichr: interactive and collaborative HTML5 gene list enrichment analysis tool. BMC Bioinformatics 14, 128 (2013).

60. Kuleshov, M. V. et al. Enrichr: a comprehensive gene set enrichment analysis web server 2016 update. Nucleic Acids Res 44, W90–97 (2016).

61. Venselaar, H., te Beek, T. A., Kuipers, R. K., Hekkelman, M. L. & Vriend, G. Protein structure analysis of mutations causing inheritable diseases. An e-Science approach with life scientist friendly interfaces. BMC Bioinformatics 11, 548 (2010).

